# Brain mechanisms explaining postural imbalance in traumatic brain injury: a systematic review

**DOI:** 10.1101/2023.07.15.23292709

**Authors:** Zaeem Hadi, Mohammad Mahmud, Barry M Seemungal

**Author notes:** Correspondence: Barry M Seemungal -; Zaeem Hadi - Centre for Vestibular Neurology, Department of Brain Sciences, Imperial College London. UK.

## Abstract

Persisting imbalance and falls in community-dwelling traumatic brain injury (TBI) survivors – typically related to vestibular dysfunction - are linked to reduced long-term survival and re-employment rates. However, a detailed understanding of the impact of TBI upon the brain mechanisms mediating imbalance is lacking. To understand the state of the art concerning the brain mechanisms mediating imbalance in TBI, we performed a systematic review of the literature.

PubMed, Web of Science, and Scopus were searched and peer-reviewed research articles in humans, with any severity of TBI (mild, moderate, severe, or concussion), that linked a postural balance assessment (objective or subjective) with brain imaging (via CT, MRI, MRS, SPECT, EEG, MEG, NIRS, and evoked potentials) were included. Out of 1940 articles, 60 were retrieved and screened, and 25 were included in the systematic review. 17 of those were MRI-based studies (e.g., DTI, fMRI), 4 EEG studies, 3 fNIRS studies, and 1 study used both MRI and EEG.

The most consistent MRI finding was the link between imbalance and cerebellum, however, the regions within the cerebellum were not consistent. Functional changes in EEG studies were non-specific as all frequency bands were reportedly linked with balance. The findings from fNIRS studies were concentrated in frontal regions as these studies only used ROI analysis. Notably, only one study reported performing clinical vestibular assessment to exclude peripheral vestibular dysfunction.

In conclusion, the lack of consistent findings could reflect that imbalance in TBI is due to a brain network dysfunction in contrast to focal cortical damage. Notably, the inconsistency in the reported findings may be attributed to heterogeneity of methodology e.g., data analytical techniques, small sample sizes, and choice of control groups. Future studies should include a detailed clinical phenotyping of vestibular function in TBI patients, ideally in an acute prospective manner, to exclude peripheral disorders. Choosing the appropriate control groups (i.e., well characterized patient subgroups as controls) would ensure that the findings are specific to imbalance rather than being non-specifically linked to TBI. Moreover, a whole-brain imaging analysis (vs ROI) is recommended to reduce selection bias and is also important since TBI affects the brain in a widespread manner.

## 1 Introduction

Traumatic brain injury (TBI) is the commonest cause of chronic disability in young adults (Langlois et al., 2006). TBI can result in vestibular dysfunction with dizziness and/or postural imbalance in up to 86% of patients acutely (Marcus et al., 2019). About 50% of TBI report imbalance at 5 years (Berman & Fredrickson, 1978) with additional socioeconomic impacts such as reduced return-to-work rates (Chamelian & Feinstein, 2004; Maskell et al., 2006). Patients are often unaware of their poor balance e.g., 50% of patients with clinically apparent gait ataxia did not report feeling unbalanced (Marcus et al., 2019; Sargeant et al., 2018; Wood et al., 2022). This can also be observed during clinical evaluation as a “vestibular agnosia”, where patients with manifest peripheral vestibular activation report little or no vertigo, and hence, vestibular dysfunction and linked imbalance in TBI could go unnoticed (Calzolari et al., 2021; Rust et al., 2022).

The mechanisms underlying balance deficits in TBI are poorly understood. The vestibular control of balance has multiple levels of vulnerability, from the vestibular organ, the nerve, brainstem and cerebellar circuits, thalamic relay areas, and cortical processing pathways. We recently demonstrated that imbalance in acute TBI was linked to the damage to corpus callosum (genu), anterior corona radiata, and external capsule (Calzolari et al., 2021). That the corpus callosum is particularly susceptible to damage in TBI because of the biomechanics of shear injury (Ghajari et al., 2017; Jolly et al., 2020), explains the observation that imbalance is virtually ubiquitous in acute TBI given its reliance upon corpus callosum integrity (Calzolari et al., 2021). Additionally, cortical processing of vestibular signals is bi-hemispheric and disruption of interhemispheric vestibular pathways (Kirsch et al., 2016) provides a means by which corpus callosal damage can impair balance.

Subcortically, vestibular afferents project to thalamus and basal ganglia (Cai et al., 2018; Stiles & Smith, 2015), with specific role of pedunculopontine nucleus (PPN) in postural balance whose neurons show high levels of vestibular reactivity (Aravamuthan & Angelaki, 2012), as implicated in the neurodegenerative diseases (i.e., Parkinson’s) (Bohnen et al., 2012; Müller et al., 2013). Indeed, Parkinson’s disease (PD) patients with severe imbalance display a vestibular agnosia and direct PPN stimulation (Yousif et al., 2016) could improve their vestibular agnosia and balance, indicating that damage or disconnection of PPN may be involved in impaired central vestibular processing resulting in impaired postural control.

Clinically well characterized TBI cohort is of critical importance for studying brain mechanisms mediating postural imbalance post-TBI. This includes the exclusion and/or treating peripheral vestibular disorders and non-specific diagnoses such as vestibular migraine. Without detailed acute clinical assessment of the TBI patients prior to neuroimaging, any measured imbalance will likely be confounded by undiagnosed and untreated peripheral disorders. A lack of established mechanism explaining postural imbalance in TBI could thus be attributed to inconsistencies in (a) detailed clinical phenotyping and excluding (or treating) patients with additional peripheral vestibular dysfunction and the secondary impact of adaptation (Conrad et al., 2022; Dieterich et al., 2007; Dieterich & Brandt, 2008; Helmchen et al., 2014) and maladaptation (Trinidade et al., 2023) upon brain correlates; (b) variation in brain imaging modality (e.g. fMRI, DTI, EEG); (c) type of analysis (e.g. whole-brain, region of interest) (Surgent et al., 2019); and (d) type of balance assessment (e.g. objective posturography, subjective scales).

Thus, we performed this systematic review to: i) synthesize the evidence linking brain imaging with imbalance in TBI to provide understanding of underlying brain mechanisms that mediate postural imbalance in TBI; and ii) identify the limitations of current literature to provide direction for future studies.

## 2 Methods

We followed PRISMA guidelines (Page et al., 2021) while performing this systematic review. The complete PRISMA checklist is provided at the end of manuscript.

### 2.1 Eligibility

Inclusion criteria for studies was: 1) **articles:** peer-reviewed research articles; 2) **participants:** humans with traumatic brain injury of any severity (concussion, mild, mild-moderate, moderate, moderate-severe, or severe) and any age group; 3) **balance assessment:** performed balance assessment (subjective scales or objective measurement (e.g. COP based measures)); 4) **brain imaging:** performed either magnetic resonance imaging (MRI, fMRI, DTI), computed tomography (CT, SPECT), positron emission tomography (PET), magnetic resonance spectroscopy (MRS), electro- or magneto-encephalography (EEG/MEG), or near-infrared spectroscopy (NIRS, fNIRS); 5) performed analysis linking balance measures (objective or subjective) with brain imaging; 6) articles in English language; 7) studies with any design are included (acute or chronic and prospective, cross-sectional, or interventional). Exclusion criteria was failing inclusion criteria, case reports, conference proceedings, research reports, and letter to the editors.

When studies used subjective symptom scales that included questions about vestibular dysfunction (including imbalance), then the study was only included if it performed an analysis linking vestibular subdomain score of the subjective scale with the brain imaging.

### 2.2 Literature Search

A literature search was conducted by first author using Pubmed, Scopus, Web of Science core collection, and Web of Science All databases (MEDLINE, Web of Science core collection, BIOSIS Citation Index, CABI: CAB Abstracts, SciELO Citation Index) from the inception of these databases until 14 March 2023. The detailed search terms for each database are provided in the appendix section.

### 2.3 Data Extraction

For the studies retrieved for detailed screening, notes were tabulated indicating inclusion or exclusion and the reason for decision by ZH, which were independently reviewed by MM. A detailed summary of the included studies was then prepared, after which the study details were tabulated. The first author (with suggestions from MM and BMS) then extracted following information from studies that were included in the systematic review: 1) study and participant characteristics; 2) scanning parameters; 3) methodological characteristics; and 4) findings reported in studies.

In terms of study and participant characteristics, we extracted information regarding first author, year of publication, study design, age, sample size, and gender for both patient and control groups. Additionally, for patients we extracted time from injury to behavioural testing, time from injury to scan, the information regarding the mode of injury (e.g., fall, traffic accident etc.), details of injury severity, and the information regarding clinical vestibular assessment and clinical scales.

For MRI studies, we also extracted information about scanning parameters including scanner vendor, magnetic field strength, number of head coil channels, type of scan (i.e., diffusion tensor imaging (DTI), resting state etc.), repetition time (TR), echo time (TE), data matrix, field of view (FOV), voxel size, and number of slices. For DTI, we further extracted information regarding diffusion directions, b-value, and number of b0 images. Moreover, number of volumes was also extracted for resting state scans. We also extracted information regarding type of imaging analysis (regions of interest (ROI), whole brain etc), balance assessments, and lastly, we extracted information about the findings showing link between brain imaging and balance assessments within TBI or comparison of TBI with controls. If multiple measures were used and reported, we extracted information for all measures and all reported findings.

For EEG/fNIRS/MEG studies, we extracted information about recording setup (electrode cap, amplifier, sampling frequency, choice of ground- and reference-electrodes, electrode locations, and the analysis software. Moreover, we also extracted information about the balance measure and the EEG/fNIRS/MEG measure used for analysis, and the electrodes that were included in the analysis.

### 2.4 Quality Assessment

We used the risk of bias assessment tool for non-randomized studies (RoBANS) (Kim et al., 2013). The studies were rated for selection bias due to inadequate selection of study participants, controlling for confounding variables, measurement of exposure, blinding of outcome assessments, incomplete outcome data (attrition bias), and reporting bias (selective reporting of outcomes). Two authors (ZH and MM) rated the studies for risk of bias and in case of disagreement, ratings were discussed and agreed upon with the senior author (BMS).

## 3 Results

### 3.1 Study Selection

Figure 1 shows the PRISMA flowchart (Haddaway et al., 2022) for the selection procedure. Our search resulted in a total of 1940 articles (415 articles from PubMed, 548 from Scopus, 429 from web of science core collection, and 548 from web of science all databases). 60 full-length articles were retrieved and assessed for eligibility. After excluding 35 articles that failed inclusion criteria, a total of 25 articles were included in this systematic review.

**Figure 1.**
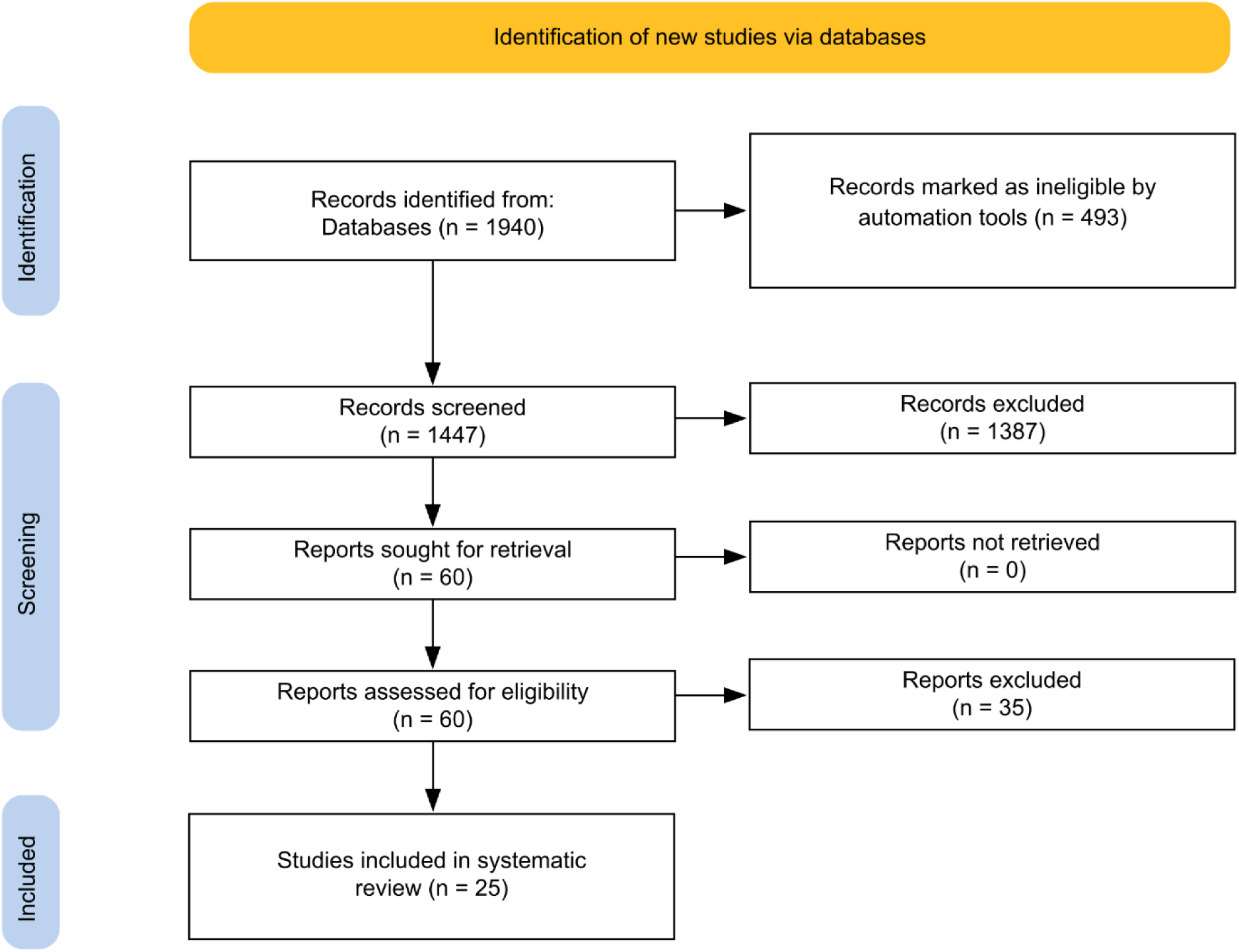
PRISMA flowchart of the selection procedure.

### 3.2 Study and Participant Characteristics

The study and participant characteristics of MRI studies are detailed in Table 1 and the clinical characteristics of injury are stated in Table 2. Importantly, due to limited EEG/fNIRS studies, and focus solely on “concussed” individuals, we decided not to compare or critically appraise EEG/fNIRS data to other imaging modalities. However, the characteristics of EEG/fNIRS studies and their findings are summarized in Table 6 and Table 7, respectively.

**Table 1.**
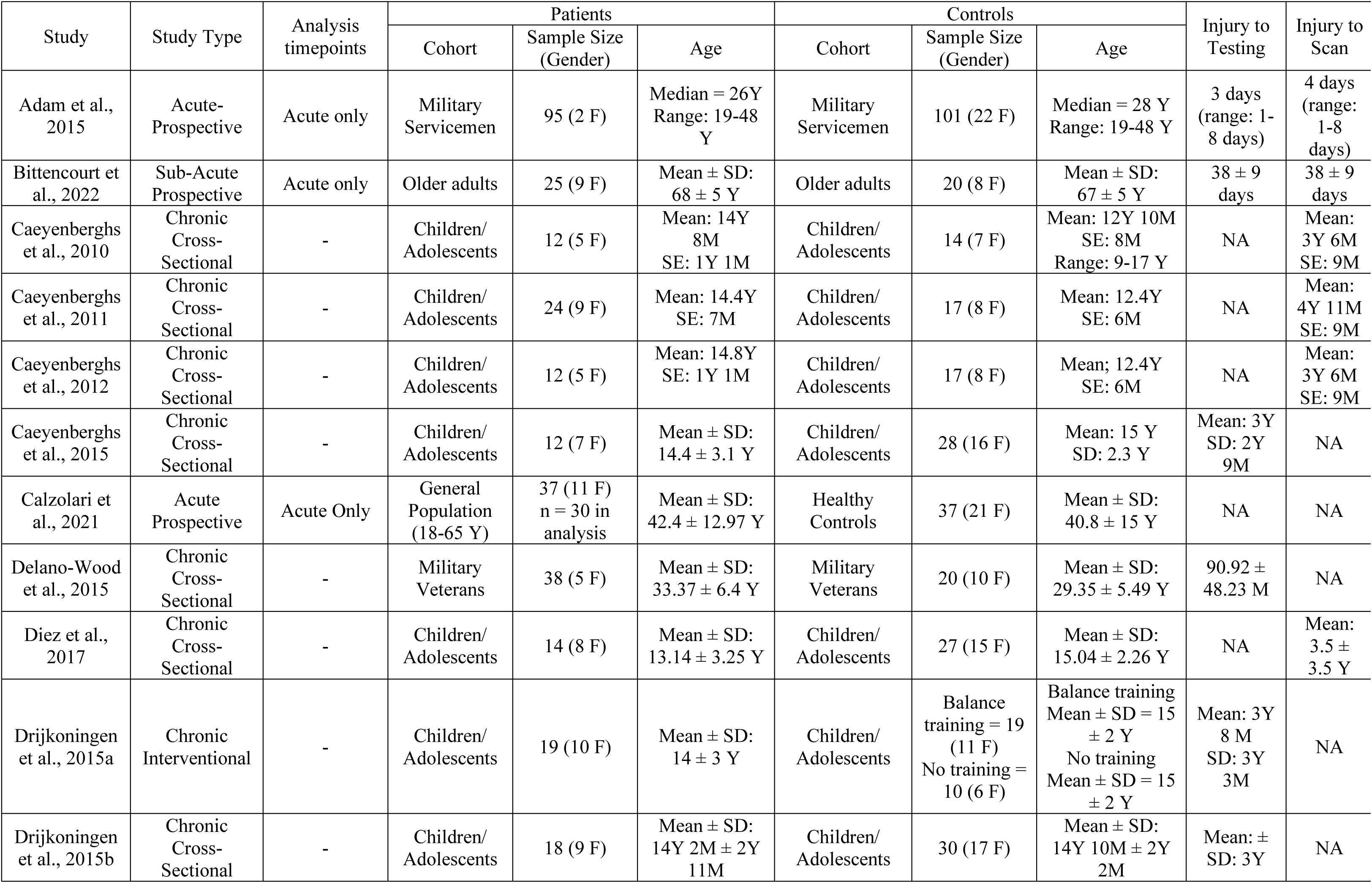
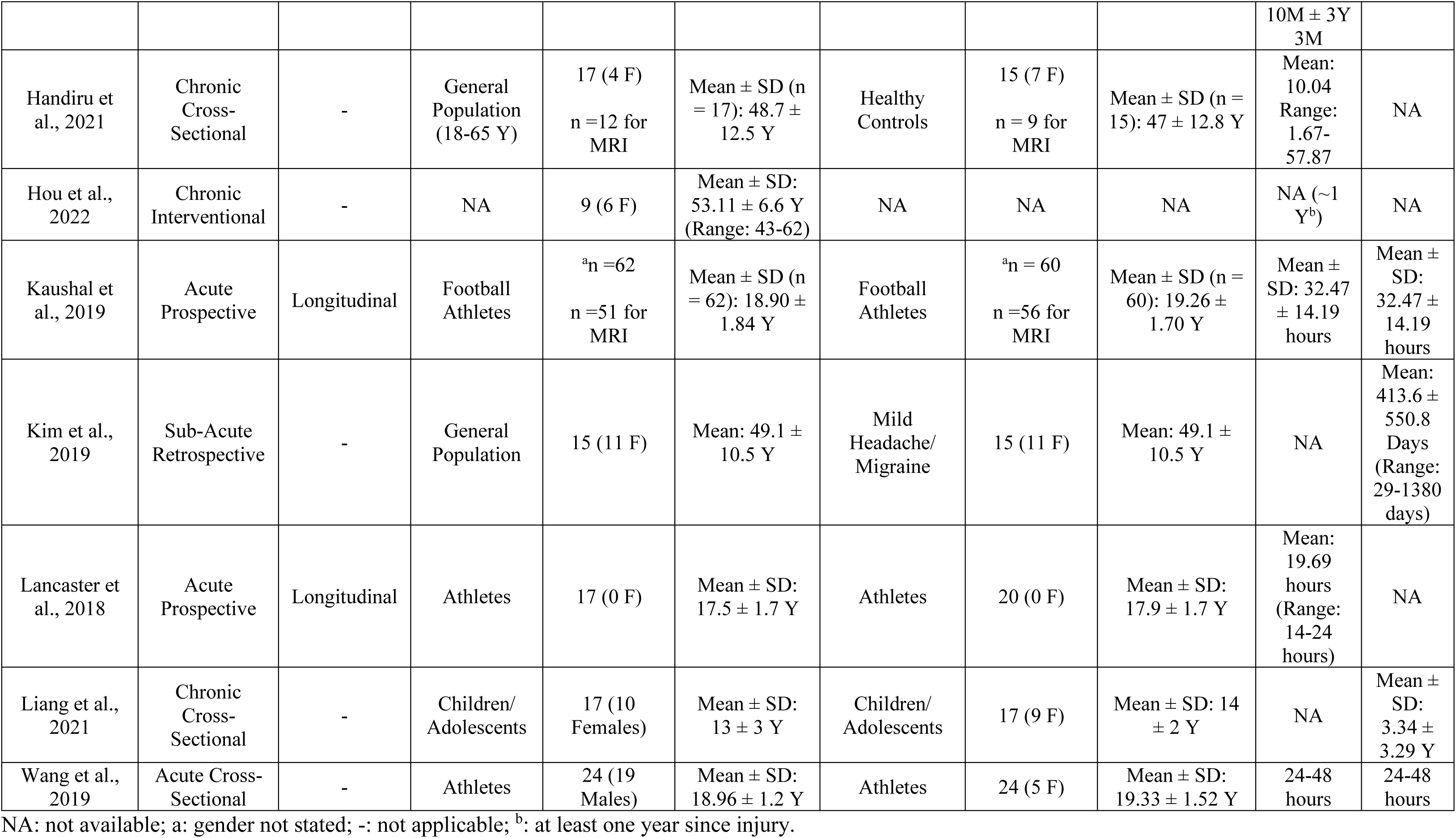
Study and Participant Characteristics.

| Study | Study Type | Analysis timepoints | Patients |  |  | Controls |  |  | Injury to Testing | Injury to Scan |
| --- | --- | --- | --- | --- | --- | --- | --- | --- | --- | --- |
|  |  |  | Cohort | Sample Size (Gender) | Age | Cohort | Sample Size (Gender) | Age |  |  |
| Adam et al., 2015 | Acute-Prospective | Acute only | Military Servicemen | 95 (2 F) | Median = 26Y<br>Range: 19-48 Y | Military Servicemen | 101 (22 F) | Median = 28 Y<br>Range: 19-48 Y | 3 days (range: 1-8 days) | 4 days (range: 1-8 days) |
| Bittencourt et al., 2022 | Sub-Acute Prospective | Acute only | Older adults | 25 (9 F) | Mean ± SD: 68 ± 5 Y | Older adults | 20 (8 F) | Mean ± SD: 67 ± 5 Y | 38 ± 9 days | 38 ± 9 days |
| Caeyenberghs et al., 2010 | Chronic Cross-Sectional | - | Children/Adolescents | 12 (5 F) | Mean: 14Y 8M<br>SE: 1Y 1M | Children/Adolescents | 14 (7 F) | Mean: 12Y 10M<br>SE: 8M<br>Range: 9-17 Y | NA | Mean: 3Y 6M<br>SE: 9M |
| Caeyenberghs et al., 2011 | Chronic Cross-Sectional | - | Children/Adolescents | 24 (9 F) | Mean: 14.4Y<br>SE: 7M | Children/Adolescents | 17 (8 F) | Mean: 12.4Y<br>SE: 6M | NA | Mean: 4Y 11M<br>SE: 9M |
| Caeyenberghs et al., 2012 | Chronic Cross-Sectional | - | Children/Adolescents | 12 (5 F) | Mean: 14.8Y<br>SE: 1Y 1M | Children/Adolescents | 17 (8 F) | Mean: 12.4Y<br>SE: 6M | NA | Mean: 3Y 6M<br>SE: 9M |
| Caeyenberghs et al., 2015 | Chronic Cross-Sectional | - | Children/Adolescents | 12 (7 F) | Mean ± SD: 14.4 ± 3.1 Y | Children/Adolescents | 28 (16 F) | Mean: 15 Y<br>SD: 2.3 Y | Mean: 3Y<br>SD: 2Y 9M | NA |
| Calzolari et al., 2021 | Acute Prospective | Acute Only | General Population (18-65 Y) | 37 (11 F)<br>n = 30 in analysis | Mean ± SD: 42.4 ± 12.97 Y | Healthy Controls | 37 (21 F) | Mean ± SD: 40.8 ± 15 Y | NA | NA |
| Delano-Wood et al., 2015 | Chronic Cross-Sectional | - | Military Veterans | 38 (5 F) | Mean ± SD: 33.37 ± 6.4 Y | Military Veterans | 20 (10 F) | Mean ± SD: 29.35 ± 5.49 Y | 90.92 ± 48.23 M | NA |
| Diez et al., 2017 | Chronic Cross-Sectional | - | Children/Adolescents | 14 (8 F) | Mean ± SD: 13.14 ± 3.25 Y | Children/Adolescents | 27 (15 F) | Mean ± SD: 15.04 ± 2.26 Y | NA | Mean: 3.5 ± 3.5 Y |
| Drijkoningen et al., 2015a | Chronic Interventional | - | Children/Adolescents | 19 (10 F) | Mean ± SD: 14 ± 3 Y | Children/Adolescents | Balance training = 19 (11 F)<br>No training = 10 (6 F) | Balance training<br>Mean ± SD = 15 ± 2 Y<br>No training<br>Mean ± SD = 15 ± 2 Y | Mean: 3Y 8 M<br>SD: 3Y 3M | NA |
| Drijkoningen et al., 2015b | Chronic Cross-Sectional | - | Children/Adolescents | 18 (9 F) | Mean ± SD: 14Y 2M ± 2Y 11M | Children/Adolescents | 30 (17 F) | Mean ± SD: 14Y 10M ± 2Y 2M | Mean: ± SD: 3Y | NA |
|  |  |  |  |  |  |  |  |  | 10M ± 3Y<br>3M |  |
| Handiru et al., 2021 | Chronic Cross-Sectional | - | General Population (18-65 Y) | 17 (4 F)<br>n =12 for MRI | Mean ± SD (n = 17): 48.7 ± 12.5 Y | Healthy Controls | 15 (7 F)<br>n = 9 for MRI | Mean ± SD (n = 15): 47 ± 12.8 Y | Mean: 10.04<br>Range: 1.67-57.87 | NA |
| Hou et al., 2022 | Chronic Interventional | - | NA | 9 (6 F) | Mean ± SD: 53.11 ± 6.6 Y (Range: 43-62) | NA | NA | NA | NA (~1 Y<sup>b</sup>) | NA |
| Kaushal et al., 2019 | Acute Prospective | Longitudinal | Football Athletes | <sup>a</sup>n =62<br>n =51 for MRI | Mean ± SD (n = 62): 18.90 ± 1.84 Y | Football Athletes | <sup>a</sup>n = 60<br>n =56 for MRI | Mean ± SD (n = 60): 19.26 ± 1.70 Y | Mean ± SD: 32.47 ± 14.19 hours | Mean ± SD: 32.47 ± 14.19 hours |
| Kim et al., 2019 | Sub-Acute Retrospective | - | General Population | 15 (11 F) | Mean: 49.1 ± 10.5 Y | Mild Headache/Migraine | 15 (11 F) | Mean: 49.1 ± 10.5 Y | NA | Mean: 413.6 ± 550.8 Days (Range: 29-1380 days) |
| Lancaster et al., 2018 | Acute Prospective | Longitudinal | Athletes | 17 (0 F) | Mean ± SD: 17.5 ± 1.7 Y | Athletes | 20 (0 F) | Mean ± SD: 17.9 ± 1.7 Y | Mean: 19.69 hours (Range: 14-24 hours) | NA |
| Liang et al., 2021 | Chronic Cross-Sectional | - | Children/Adolescents | 17 (10 Females) | Mean ± SD: 13 ± 3 Y | Children/Adolescents | 17 (9 F) | Mean ± SD: 14 ± 2 Y | NA | Mean ± SD: 3.34 ± 3.29 Y |
| Wang et al., 2019 | Acute Cross-Sectional | - | Athletes | 24 (19 Males) | Mean ± SD: 18.96 ± 1.2 Y | Athletes | 24 (5 F) | Mean ± SD: 19.33 ± 1.52 Y | 24-48 hours | 24-48 hours |

NA: not available; a: gender not stated; -: not applicable; <sup>b</sup>: at least one year since injury.

**Table 2.**
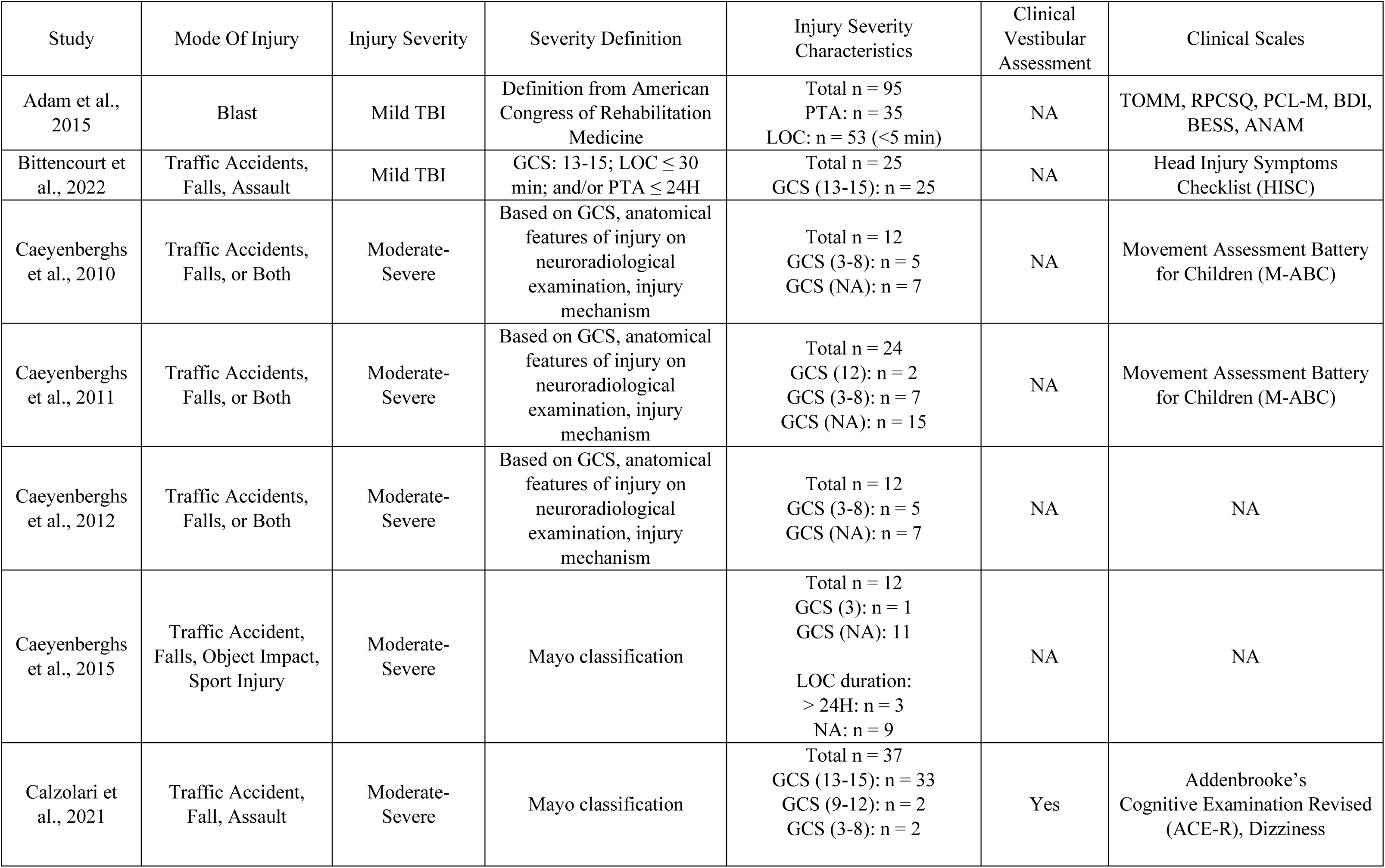
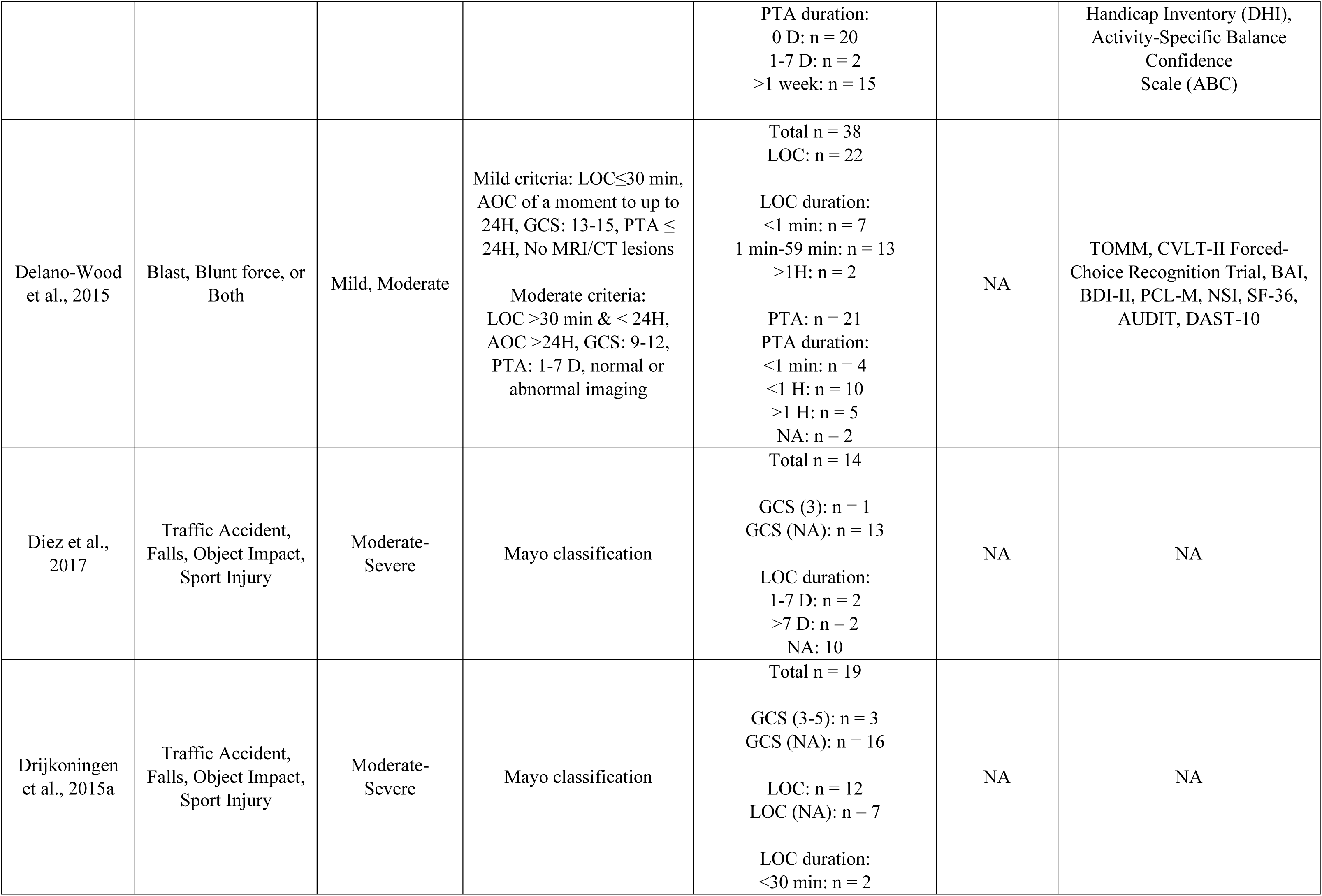
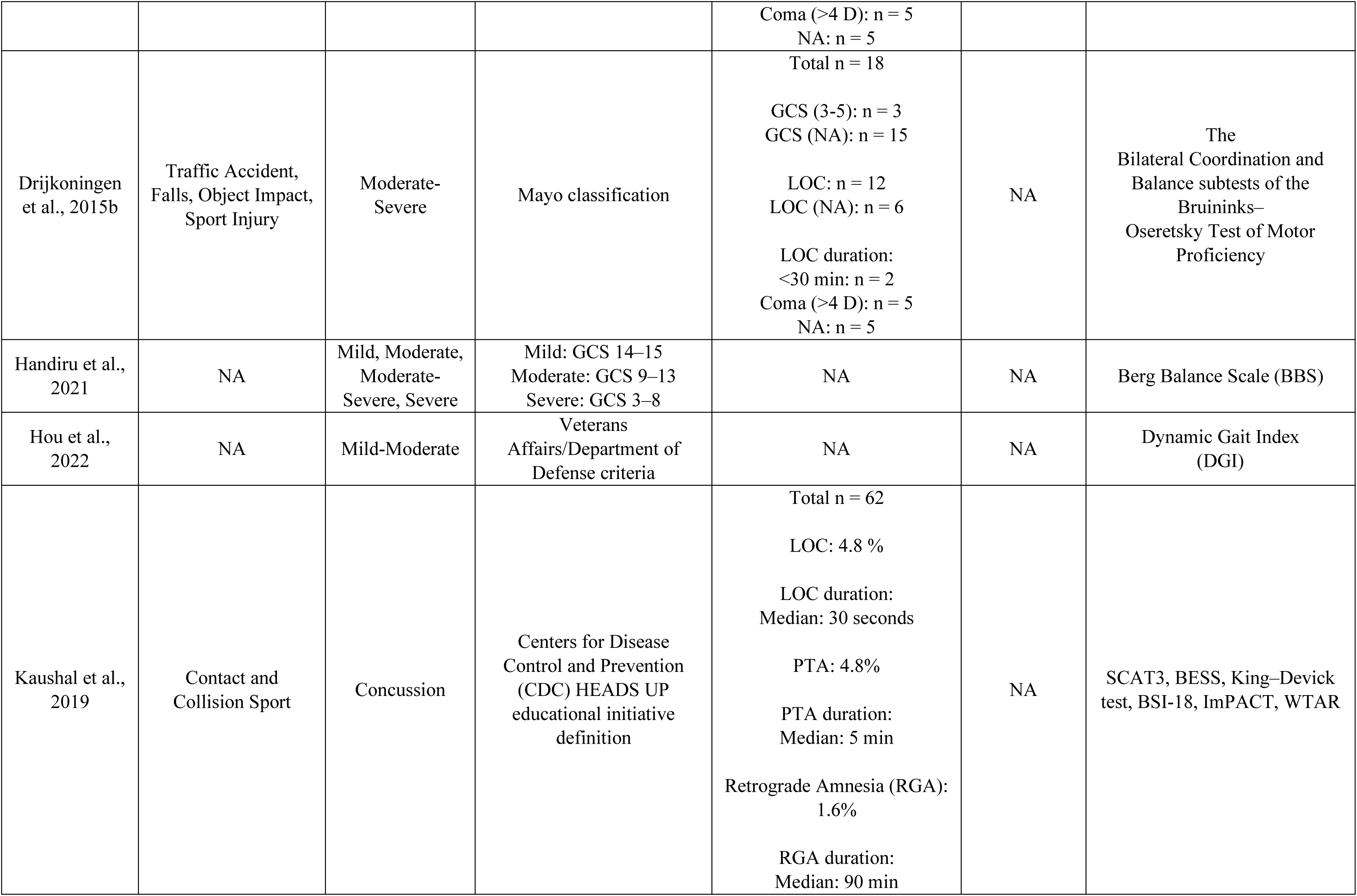
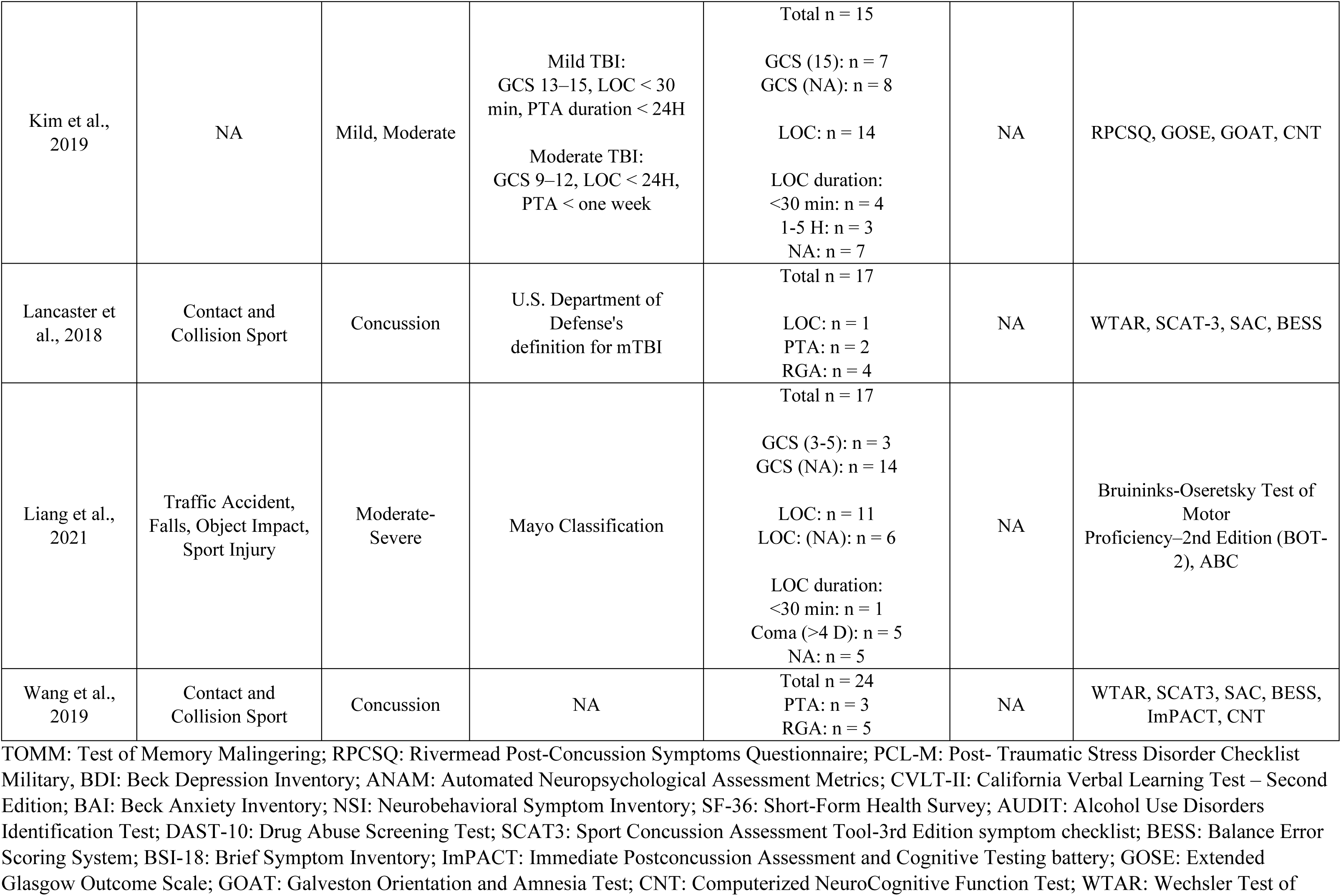
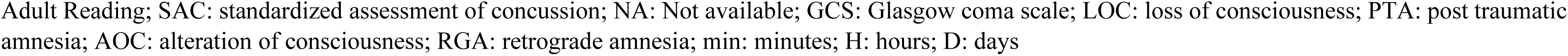
Participant injury and clinical characteristics.

| Study | Mode Of Injury | Injury Severity | Severity Definition | Injury Severity Characteristics | Clinical Vestibular Assessment | Clinical Scales |
| --- | --- | --- | --- | --- | --- | --- |
| Adam et al., 2015 | Blast | Mild TBI | Definition from American Congress of Rehabilitation Medicine | Total n = 95<br>PTA: n = 35<br>LOC: n = 53 (<5 min) | NA | TOMM, RPCSQ, PCL-M, BDI, BESS, ANAM |
| Bittencourt et al., 2022 | Traffic Accidents, Falls, Assault | Mild TBI | GCS: 13-15; LOC ≤ 30 min; and/or PTA ≤ 24H | Total n = 25<br>GCS (13-15): n = 25 | NA | Head Injury Symptoms Checklist (HISC) |
| Caeyenberghs et al., 2010 | Traffic Accidents, Falls, or Both | Moderate-Severe | Based on GCS, anatomical features of injury on neuroradiological examination, injury mechanism | Total n = 12<br>GCS (3-8): n = 5<br>GCS (NA): n = 7 | NA | Movement Assessment Battery for Children (M-ABC) |
| Caeyenberghs et al., 2011 | Traffic Accidents, Falls, or Both | Moderate-Severe | Based on GCS, anatomical features of injury on neuroradiological examination, injury mechanism | Total n = 24<br>GCS (12): n = 2<br>GCS (3-8): n = 7<br>GCS (NA): n = 15 | NA | Movement Assessment Battery for Children (M-ABC) |
| Caeyenberghs et al., 2012 | Traffic Accidents, Falls, or Both | Moderate-Severe | Based on GCS, anatomical features of injury on neuroradiological examination, injury mechanism | Total n = 12<br>GCS (3-8): n = 5<br>GCS (NA): n = 7 | NA | NA |
| Caeyenberghs et al., 2015 | Traffic Accident, Falls, Object Impact, Sport Injury | Moderate-Severe | Mayo classification | Total n = 12<br>GCS (3): n = 1<br>GCS (NA): 11<br>LOC duration:<br>> 24H: n = 3<br>NA: n = 9 | NA | NA |
| Calzolari et al., 2021 | Traffic Accident, Fall, Assault | Moderate-Severe | Mayo classification | Total n = 37<br>GCS (13-15): n = 33<br>GCS (9-12): n = 2<br>GCS (3-8): n = 2 | Yes | Addenbrooke's Cognitive Examination Revised (ACE-R), Dizziness |
|  |  |  |  | PTA duration:<br>0 D: n = 20<br>1-7 D: n = 2<br>>1 week: n = 15 |  | Handicap Inventory (DHI),<br>Activity-Specific Balance<br>Confidence<br>Scale (ABC) |
| Delano-Wood<br>et al., 2015 | Blast, Blunt force, or<br>Both | Mild, Moderate | Mild criteria: LOC ≤ 30 min,<br>AOC of a moment to up to<br>24H, GCS: 13-15, PTA ≤<br>24H, No MRI/CT lesions<br>Moderate criteria:<br>LOC > 30 min & < 24H,<br>AOC > 24H, GCS: 9-12,<br>PTA: 1-7 D, normal or<br>abnormal imaging | Total n = 38<br>LOC: n = 22<br>LOC duration:<br><1 min: n = 7<br>1 min-59 min: n = 13<br>>1H: n = 2<br>PTA: n = 21<br>PTA duration:<br><1 min: n = 4<br><1 H: n = 10<br>>1 H: n = 5<br>NA: n = 2 | NA | TOMM, CVLT-II Forced-Choice Recognition Trial, BAI,<br>BDI-II, PCL-M, NSI, SF-36,<br>AUDIT, DAST-10 |
| Diez et al.,<br>2017 | Traffic Accident,<br>Falls, Object Impact,<br>Sport Injury | Moderate-Severe | Mayo classification | Total n = 14<br>GCS (3): n = 1<br>GCS (NA): n = 13<br>LOC duration:<br>1-7 D: n = 2<br>>7 D: n = 2<br>NA: 10 | NA | NA |
| Drijkoningen<br>et al., 2015a | Traffic Accident,<br>Falls, Object Impact,<br>Sport Injury | Moderate-Severe | Mayo classification | Total n = 19<br>GCS (3-5): n = 3<br>GCS (NA): n = 16<br>LOC: n = 12<br>LOC (NA): n = 7<br>LOC duration:<br><30 min: n = 2 | NA | NA |
|  |  |  |  | Coma (>4 D): n = 5<br>NA: n = 5 |  |  |
| Drijkoningen<br>et al., 2015b | Traffic Accident,<br>Falls, Object Impact,<br>Sport Injury | Moderate-Severe | Mayo classification | Total n = 18<br>GCS (3-5): n = 3<br>GCS (NA): n = 15<br>LOC: n = 12<br>LOC (NA): n = 6<br>LOC duration:<br><30 min: n = 2<br>Coma (>4 D): n = 5<br>NA: n = 5 | NA | The<br>Bilateral Coordination and<br>Balance subtests of the<br>Bruininks–<br>Oseretsky Test of Motor<br>Proficiency |
| Handiru et al.,<br>2021 | NA | Mild, Moderate,<br>Moderate-Severe, Severe | Mild: GCS 14–15<br>Moderate: GCS 9–13<br>Severe: GCS 3–8 | NA | NA | Berg Balance Scale (BBS) |
| Hou et al.,<br>2022 | NA | Mild-Moderate | Veterans<br>Affairs/Department of<br>Defense criteria | NA | NA | Dynamic Gait Index<br>(DGI) |
| Kaushal et al.,<br>2019 | Contact and<br>Collision Sport | Concussion | Centers for Disease<br>Control and Prevention<br>(CDC) HEADS UP<br>educational initiative<br>definition | Total n = 62<br>LOC: 4.8 %<br>LOC duration:<br>Median: 30 seconds<br>PTA: 4.8%<br>PTA duration:<br>Median: 5 min<br>Retrograde Amnesia (RGA):<br>1.6%<br>RGA duration:<br>Median: 90 min | NA | SCAT3, BESS, King–Devick<br>test, BSI-18, ImPACT, WTAR |
| Kim et al.,<br>2019 | NA | Mild, Moderate | Mild TBI:<br>GCS 13–15, LOC < 30<br>min, PTA duration < 24H<br>Moderate TBI:<br>GCS 9–12, LOC < 24H,<br>PTA < one week | Total n = 15<br>GCS (15): n = 7<br>GCS (NA): n = 8<br>LOC: n = 14<br>LOC duration:<br><30 min: n = 4<br>1-5 H: n = 3<br>NA: n = 7 | NA | RPCSQ, GOSE, GOAT, CNT |
| Lancaster et<br>al., 2018 | Contact and<br>Collision Sport | Concussion | U.S. Department of<br>Defense's<br>definition for mTBI | Total n = 17<br>LOC: n = 1<br>PTA: n = 2<br>RGA: n = 4 | NA | WTAR, SCAT-3, SAC, BESS |
| Liang et al.,<br>2021 | Traffic Accident,<br>Falls, Object Impact,<br>Sport Injury | Moderate-Severe | Mayo Classification | Total n = 17<br>GCS (3-5): n = 3<br>GCS (NA): n = 14<br>LOC: n = 11<br>LOC: (NA): n = 6<br>LOC duration:<br><30 min: n = 1<br>Coma (>4 D): n = 5<br>NA: n = 5 | NA | Bruininks-Oseretsky Test of<br>Motor<br>Proficiency–2nd Edition (BOT-2), ABC |
| Wang et al.,<br>2019 | Contact and<br>Collision Sport | Concussion | NA | Total n = 24<br>PTA: n = 3<br>RGA: n = 5 | NA | WTAR, SCAT3, SAC, BESS,<br>ImPACT, CNT |

TOMM: Test of Memory Malinger; RPCSQ: Rivermead Post-Concussion Symptoms Questionnaire; PCL-M: Post- Traumatic Stress Disorder Checklist Military, BDI: Beck Depression Inventory; ANAM: Automated Neuropsychological Assessment Metrics; CVLT-II: California Verbal Learning Test – Second Edition; BAI: Beck Anxiety Inventory; NSI: Neurobehavioral Symptom Inventory; SF-36: Short-Form Health Survey; AUDIT: Alcohol Use Disorders Identification Test; DAST-10: Drug Abuse Screening Test; SCAT3: Sport Concussion Assessment Tool-3rd Edition symptom checklist; BESS: Balance Error Scoring System; BSI-18: Brief Symptom Inventory; ImPACT: Immediate Postconcussion Assessment and Cognitive Testing battery; GOSE: Extended Glasgow Outcome Scale; GOAT: Galveston Orientation and Amnesia Test; CNT: Computerized NeuroCognitive Function Test; WTAR: Wechsler Test of
Adult Reading; SAC: standardized assessment of concussion; NA: Not available; GCS: Glasgow coma scale; LOC: loss of consciousness; PTA: post traumatic amnesia; AOC: alteration of consciousness; RGA: retrograde amnesia; min: minutes; H: hours; D: days

It is important to highlight that there was only one MRI acute-prospective study of moderate-severe TBI (Calzolari et al., 2021). Moreover, there were only two studies in the general population of age 18-65 (Calzolari et al., 2021; Handiru et al., 2021). Most of the other studies (n = 8) were in young athletes or children/adolescents. Since most of the studies performed comparisons with control group (between-group analysis), the sample sizes of studies were considered small to medium with only two studies having relatively larger sample of patients n = 95 (Adam et al., 2015) and n = 62 (Kaushal et al., 2019), where n = 51 were included in MRI analysis in (Kaushal et al., 2019). Most importantly, peripheral vestibular dysfunction (i.e., problem with vestibular organ or vestibular nerve) or other vestibular diagnoses e.g., vestibular migraine, were not ruled out in any of the studies nor was it mentioned as exclusion criteria, except in one study (Calzolari et al., 2021).

Three MRI studies used a partially overlapping patient cohort (Caeyenberghs et al., 2010, 2011, 2012), and five other MRI studies also used a partially overlapping patient cohort (Caeyenberghs et al., 2015; Diez et al., 2017; Drijkoningen et al., 2015a; Drijkoningen et al., 2015b; Liang et al., 2021). This is based on a detailed comparison of participant information, and injury details stated in these articles (i.e., demographics, GCS score, injury specifics on MRI and CT). Only one study (Liang et al., 2021) explicitly declared the use of data collected and published previously (Drijkoningen et al., 2015a).

### 3.3 Quality Assessment

#### Selection of participants

Methodological quality of the studies is visualized in Figure 2 using *robvis* tool (McGuinness & Higgins, 2021). Two studies (Handiru et al., 2021; Kim et al., 2019) included TBI patients with mild and moderate or higher severity in same analysis; moreover, mode of injury (i.e., fall, traffic accident etc.) for patients is also not available in both studies. Kim et al., 2019 included 7 moderate and 8 mild TBI in the brain imaging analyses and also also reported to not have obtained informed consent of study participants due to retrospective nature of the study. Handiru et al., 2021 originally had a sample of n =17 (3 mild, 3 moderate, 3 moderate/severe (diagnosis not confirmed), and 8 severe TBI), however, only n = 12 were included in the imaging analysis. The proportions of mild, moderate, or severe TBI in this sample of n = 12 were not stated in the study. Given that moderate-severe TBI patients’ structural imaging analysis is more likely to influence the findings as compared to mild TBI and for reasons listed above, we decided to rate (Handiru et al., 2021; Kim et al., 2019) as “high risk” for selection of participants. (Kaushal et al., 2019) did not specify the gender of study participants and was thus rated as “unclear risk” on selection of study participants. While (Delano-Wood et al., 2015) combined the analysis of moderate TBI patients (n = 5) and mild TBI (n = 33), they report that the inclusion or removal of the moderate severity TBI did not affect the results. Calzolari et al., 2021 had 4 participants with mild-probable TBI severity in a sample of n = 37, however, a subsample of n = 30 of 37 was included in imaging analysis such that only 2 mild/probable TBI were included in analysis, and thus we considered it to be a “low risk”.

**Figure 2.**
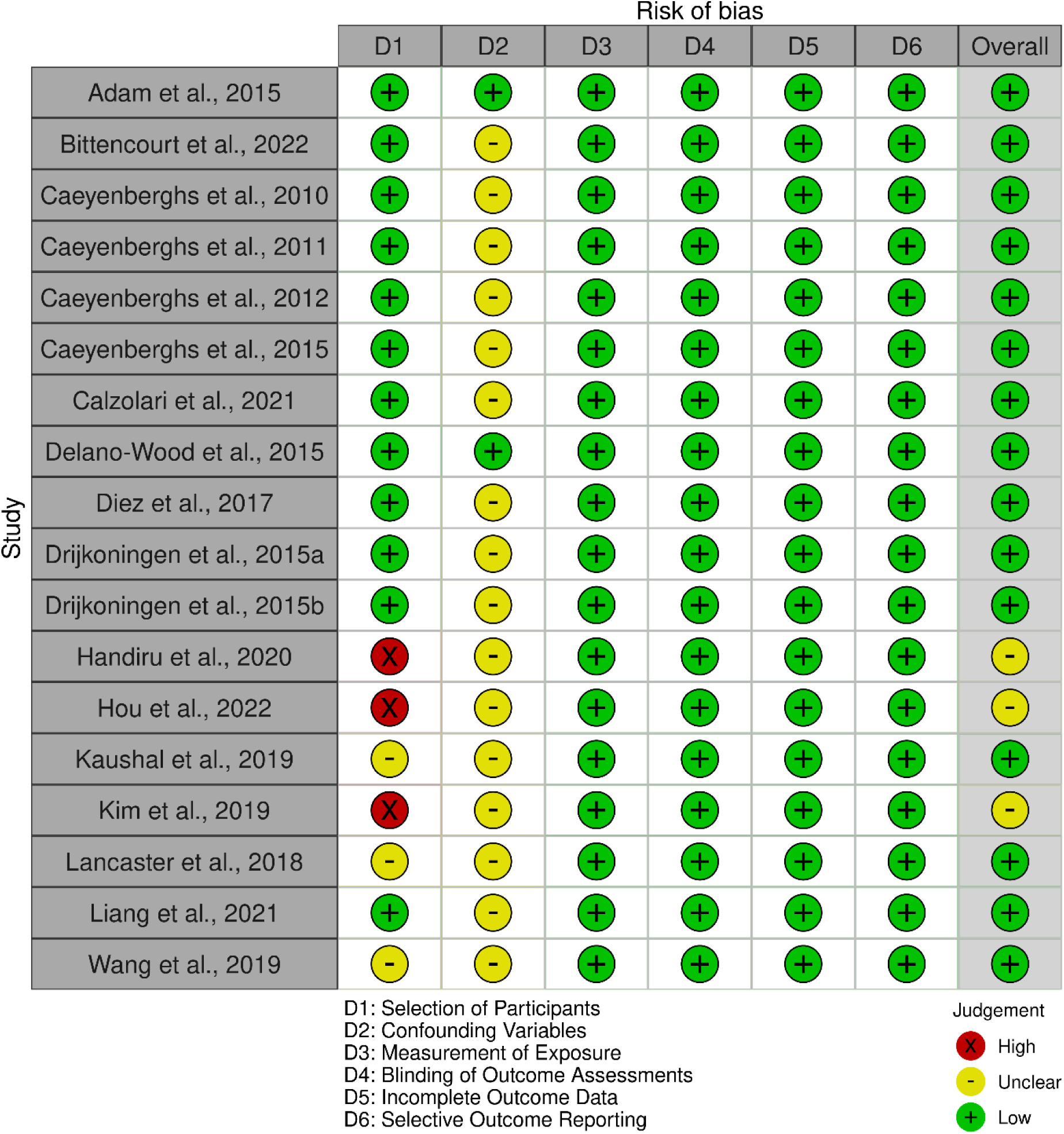
Quality Assessment of Studies using ROBANS risk of bias assessment tool.

Hou et al., 2022 used US departments of defense (DoD) and veteran affairs (VA) criteria to classify patients as “mild to moderate” TBI. Notably, DOD/VA criteria differentiates between “mild” and “moderate” but does not specify a category label of “mild to moderate”. Thus, the proportions of mild or moderate TBI patients were not explicitly stated in the study (Hou et al., 2022). Moreover, as the study was an interventional study, there was no control group to assess whether the pre-post intervention change of balance outcome in the patients is not due to test-retest variability. Thus, due to lack of information regarding injury classification and the lack of appropriate control group, we rated Hou et al., 2022 as “high risk” on selection of participants.

Since mild/concussed TBI patients, as classified based on absence of CT abnormalities, could still have MRI abnormalities, the studies with mild and concussed athletes were also rated as “unclear risk” in case: i) if they did not explicitly state exclusion of participants with a lesion or incidental MRI finding; or ii) if the mTBI/concussion definition did not explicitly address exclusion of imaging findings. Lancaster et al., 2018 used the US DOD definition of “mild” TBI to define sports related concussion. Notably, US DOD definition requires normal brain imaging for participants to be considered mild TBI, however, the definition stated in Lancaster et al., 2018 does not include any mention of normal brain imaging as inclusion criteria. Similarly, Wang et al., 2019 stated no details in study regarding the definition of injury or concussion. Thus, both of these studies were rated as “unclear risk” on selection of participants.

#### Control for confounding variables

In reference to accounting for confounding variables, none of the studies mapped the lesions in their analysis. Adam et al., 2015 stated that no MRI abnormalities were detected in participants whereas Liang et al. 2021 explicitly addressed the issue of lesion mapping and given the heterogenous lesions among their participants, they chose to not perform lesion masking. Similarly, Calzolari et al., 2021 chose not to perform lesion mapping since some of the severe TBI patients did not show lesions. Since lesion volume could instead be used as a confound regressor (given the heterogeneity of lesions in TBI), all studies that did not account for lesions or did not explicitly address the issue of lesion mapping were rated as “unclear risk” for confounding variables.

### 3.4 Scanning Characteristics

The details of scanning parameters of MRI studies are listed in Table 3. All studies used a 3T scanner except one study (Adam et al., 2015) that used 1.5T scanner. There was one study (Wang et al., 2019) that used arterial spin labelling (ASL), which is an fMRI technique, and also had two different study sites with 4 mm and 4.5 mm slice thickness. Notably, Wang et al., 2019 and Kaushal et al., 2019 only reported slice thickness but did not report voxel sizes of the scans acquired. Two resting-state studies (Bittencourt et al., 2022; Hou et al., 2022) also had voxel size of 3.5 mm (isotropic) that were relatively larger than other studies.

**Table 3.** Scanning Characteristics.

| Study | Scanner | Magnetic field strength | Head coil channels | Imaging Modality | Diffusion directions | b-value (s/mm<sup>2</sup>) | No. of b0 | Repetition time (ms) - TR | Echo time (ms) - TE | Data Matrix | FOV (mm<sup>2</sup>) | Voxel Size (mm) | No. of Slices | Volumes |
| --- | --- | --- | --- | --- | --- | --- | --- | --- | --- | --- | --- | --- | --- | --- |
| Adam et al., 2015 | Philips Achieva | 1.5 T | NA | DTI | 15 | 1000 | 1 | NA | NA | NA | NA | 2.5 isotropic | NA | - |
| Bittencourt et al., 2022 | Siemens Magnetom Prisma | 3 T | 64 | Resting state | NA | NA | NA | 2000 | 9.74, 22.10, 34.46 | NA | 256 × 256 | 3.5 isotropic | NA | 300 |
| Caeyenberghs et al., 2010 | Philips Intera | 3 T | 8 | DTI | 45 | 800 | 1 | 7916 | 68 | 112 x 112 | 220 × 220 | 2 × 2 × 2.2 | 68 | - |
| Caeyenberghs et al., 2011 | Philips Intera | 3 T | 8 | DTI | 45 | 800 | 1 | 7916 | 68 | 112 x 112 | 220 × 220 | 2 × 2 × 2.2 | 68 | - |
| Caeyenberghs et al., 2012 | Philips Intera | 3 T | 8 | DTI | 45 | 800 | 1 | 7916 | 68 | 112 x 112 | 220 × 220 | 2 × 2 × 2.2 | 68 | - |
| Caeyenberghs et al., 2015 | Siemens Magnetom Trio | 3 T | 12 | Resting State | - | - | - | 3000 | 30 | 80 x 80 | 230 × 230 | 2.5 × 2.5 × 3.1 Slice thickness: 2.8 | 50 | 200 |
| Calzolari et al., 2021 | Siemens Verio | 3 T | 32 | DTI | 64 | 1000 | 4 | 9500 | 103 | 128 x 128 | 256 × 256 | 2 isotropic | 64 | - |
| Delano-Wood et al., 2015 | General Electric MR750w | 3 T | 8 | DTI | 25 | 1500 | 1 | 8000 | Range: 81.2-86 | 128 x 128 | 240 | 1.875 × 1.875 Slice thickness: 1.5 | 64 | - |
| Diez et al., 2017 | Siemens Magnetom Trio | 3 T | 12 | DTI | 64 | 1000 | NA | 8000 | 91 | NA | 212 × 212 | 2.2 isotropic | 60 | - |
|  |  |  |  | Resting State | - | - | - | 3000 | 30 | 80 x 80 | 230 × 230 | 2.5 × 2.5 × 3.1 Slice thickness: 2.8 | 50 | 200 |
| Drijkoningen et al., 2015a | Siemens Magnetom Trio | 3 T | 12 | DTI | 64 | 1000 | 1 | 8000 | 91 | NA | 212 × 212 | 2.2 isotropic | 60 | - |
| Drijkoningen et al., 2015b | Siemens Magnetom Trio | 3 T | 12 | T1 (VBM) | - | - | - | 2300 | 2.98 | NA | 256 × 240 | 1 × 1 × 1.1 | 160 | - |
| Handiru et al., 2021 | Siemens Skyra | 3 T | NA | DTI | 64 | 1100 | 8 | 9000 | 75 | 128 x 128 | 256 × 256 | 2 isotropic | 66 | - |
| Hou et al., 2022 | General Electric GE750 | 3 T | NA | Resting State | - | - | - | 2000 | 22 | NA | 100 × 100 | 3.5 isotropic | NA | NA |
| Kaushal et al., 2019 | General Electric MR750 | 3 T | 32 | Resting State | - | - | - | 720 | 30 | 104 x 104 | 210 | <sup>a</sup>Slice thickness: 2 | 72 | 501 |
| Kim et al., 2019 | General Electric MR750w Discovery | 3 T | NA | DTI | 15 | 1000 | 1 | 8000 | Range: 76.1 - 85 | 128 x 128 | 240 | 0.9 × 0.9 × 4 Slice thickness: 2 | Range: 34-37 | - |
| Lancaster et al., 2018 | General Electric MR750w Discovery | 3 T | 32 | DTI | 30 | 1000 & 2000 | 5 (1 in opposite phase) | 5250 | 66.8 | NA | NA | 3 isotropic | NA | - |
| Liang et al., 2021 | Siemens Trio | 3 T | 12 | DTI | 64 | 1000 | 1 | 8000 | 91 | NA | NA | 2.2 isotropic | 60 | - |
| Wang et al., 2019 | Site 1: Siemens Trio | 3 T | 32 | ASL | - | - | - | 3204 | TE: 13<br>TI<sub>1</sub>: 700<br>TI<sub>1S</sub>: 1600<br>TI<sub>2</sub>: 1800 | 64 × 64 | 224 | <sup>a</sup>Slice thickness: 4.5 | 36 | 109 (54 Control and label image pairs)<br>One M<sub>0</sub> image |
|  | Site 2: General Electric Discovery MR750w | 3 T | 32 | ASL | - | - | - | 4632 | TE: 10.5<br>TI: NA | 128 × 128 | 240 | <sup>a</sup>Slice thickness: 4 | 36 | - |

-: not applicable; NA: Not available; T: tesla; DTI: diffusion tensor imaging; ASL: arterial spin labelling; PA: posterior-anterior. <sup>a</sup>: voxel size not provided

### 3.5 Methodological Characteristics

#### Balance assessments

The measure used for balance assessment, type of analyses used for analysing MRI images, and the imaging metric used for correlating with balance measure are listed in Table 4. Seven studies used subjective balance scales, of which four studies used BESS (Adam et al., 2015; Kaushal et al., 2019; Lancaster et al., 2018; Wang et al., 2019), one study used M-ABC (Caeyenberghs et al., 2011), one study used NSI-vestibular subscale (Delano-Wood et al., 2015), and one study used HISC (vestibular sub-domain) (Bittencourt et al., 2022). One study (Handiru et al., 2021) used both, a subjective (Berg balance scale – BBS) and objective balance assessment (posturography).

**Table 4.**
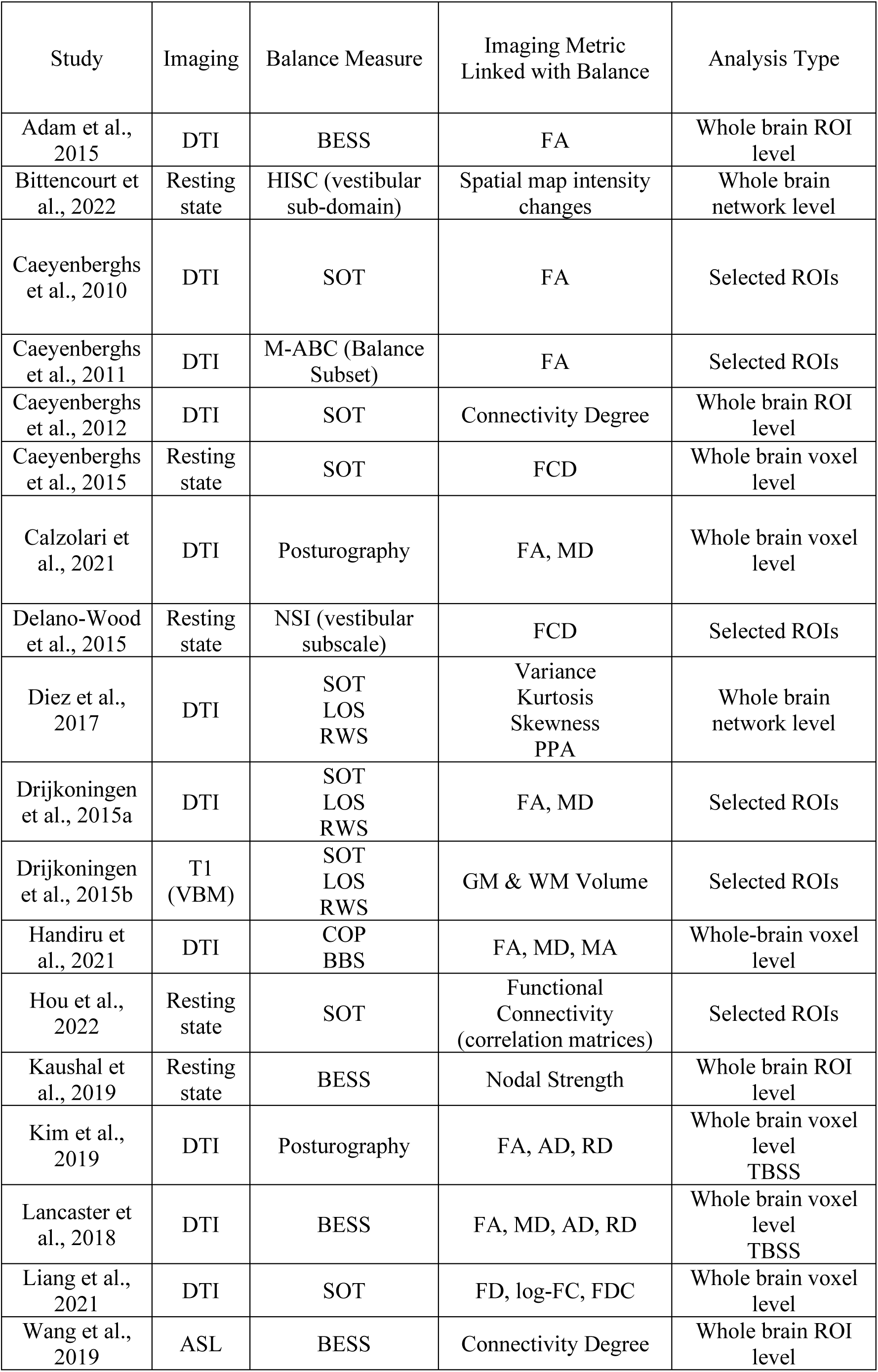
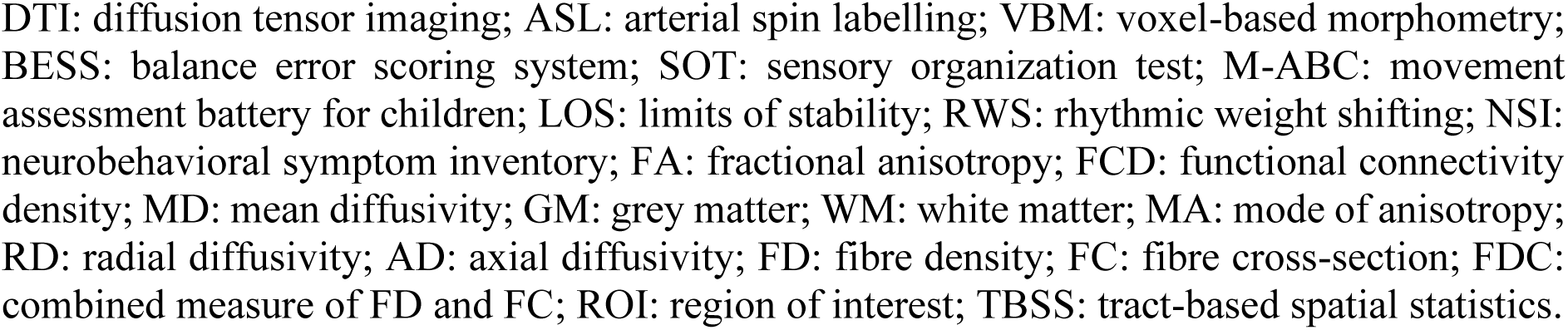
Analysis Methods and Balance Assessment in Included Studies.

| Study | Imaging | Balance Measure | Imaging Metric Linked with Balance | Analysis Type |
| --- | --- | --- | --- | --- |
| Adam et al., 2015 | DTI | BESS | FA | Whole brain ROI level |
| Bittencourt et al., 2022 | Resting state | HISC (vestibular sub-domain) | Spatial map intensity changes | Whole brain network level |
| Caeyenberghs et al., 2010 | DTI | SOT | FA | Selected ROIs |
| Caeyenberghs et al., 2011 | DTI | M-ABC (Balance Subset) | FA | Selected ROIs |
| Caeyenberghs et al., 2012 | DTI | SOT | Connectivity Degree | Whole brain ROI level |
| Caeyenberghs et al., 2015 | Resting state | SOT | FCD | Whole brain voxel level |
| Calzolari et al., 2021 | DTI | Posturography | FA, MD | Whole brain voxel level |
| Delano-Wood et al., 2015 | Resting state | NSI (vestibular subscale) | FCD | Selected ROIs |
| Diez et al., 2017 | DTI | SOT<br>LOS<br>RWS | Variance<br>Kurtosis<br>Skewness<br>PPA | Whole brain network level |
| Drijkoningen et al., 2015a | DTI | SOT<br>LOS<br>RWS | FA, MD | Selected ROIs |
| Drijkoningen et al., 2015b | T1 (VBM) | SOT<br>LOS<br>RWS | GM & WM Volume | Selected ROIs |
| Handiru et al., 2021 | DTI | COP<br>BBS | FA, MD, MA | Whole-brain voxel level |
| Hou et al., 2022 | Resting state | SOT | Functional Connectivity (correlation matrices) | Selected ROIs |
| Kaushal et al., 2019 | Resting state | BESS | Nodal Strength | Whole brain ROI level |
| Kim et al., 2019 | DTI | Posturography | FA, AD, RD | Whole brain voxel level<br>TBSS |
| Lancaster et al., 2018 | DTI | BESS | FA, MD, AD, RD | Whole brain voxel level<br>TBSS |
| Liang et al., 2021 | DTI | SOT | FD, log-FC, FDC | Whole brain voxel level |
| Wang et al., 2019 | ASL | BESS | Connectivity Degree | Whole brain ROI level |

DTI: diffusion tensor imaging; ASL: arterial spin labelling; VBM: voxel-based morphometry; BESS: balance error scoring system; SOT: sensory organization test; M-ABC: movement assessment battery for children; LOS: limits of stability; RWS: rhythmic weight shifting; NSI: neurobehavioral symptom inventory; FA: fractional anisotropy; FCD: functional connectivity density; MD: mean diffusivity; GM: grey matter; WM: white matter; MA: mode of anisotropy; RD: radial diffusivity; AD: axial diffusivity; FD: fibre density; FC: fibre cross-section; FDC: combined measure of FD and FC; ROI: region of interest; TBSS: tract-based spatial statistics.

Nine studies used objective balance assessments, of which five studies used sensory organization test (SOT) (Caeyenberghs et al., 2010, 2012, 2015; Hou et al., 2022; Liang et al., 2021), two studies used posturography (Calzolari et al., 2021; Kim et al., 2019), and three studies used SOT, limits of stability (LOS) test, and rhythmic weight shift (RWS) test, together (Diez et al., 2017; Drijkoningen et al., 2015a; Drijkoningen et al., 2015b).

#### Types of brain imaging analyses

Six studies used selected ROIs for the analysis (Caeyenberghs et al., 2010, 2011; Delano-Wood et al., 2015; Drijkoningen et al., 2015a; Drijkoningen et al., 2015b; Hou et al., 2022). Caeyenberghs et al., 2010 used nine subcortical white-matter tracts/regions as ROIs including superior-middle-inferior cerebellar peduncles (SCP, MCP, ICP), cerebellum, brainstem, pons, corticospinal tract (CST), medial lemniscus (ML), cerebral peduncles (CP), and three cortical white-matter tracts anterior-posterior limb of the internal capsule (ALIC, PLIC), and posterior thalamic radiation (PTR). Caeyenberghs et al., 2011 used seven sub-cortical ROIs including CST, SCP, ICP, CP, thalamus, cerebellum, brain stem, and four cortical ROIs including corpus callosum (CC), anterior corona radiata (ACR), ALIC, and PLIC. Drijkoningen et al., 2015a mainly focused on subcortical ROIs including SCP, MCP, ICP, and cerebellum. Similarly, Drijkoningen et al., 2015b also only focused on subcortical ROIs including cerebellum and brain stem. Delano-Wood et al., 2015 also used subcortical tracts as ROIs including CST, pontine tegmentum, ML, and central tegmental tract. Hou et al., 2022 used 35 ROIs from sensory/somatomotor resting state network atlas, 31 ROIs from visual network, and 4 ROIs from cerebellar network.

Four studies performed whole-brain analysis at ROI level (Adam et al., 2015; Caeyenberghs et al., 2012; Kaushal et al., 2019; Wang et al., 2019) and six studies performed whole-brain voxel level analysis (Caeyenberghs et al., 2015; Calzolari et al., 2021; Handiru et al., 2021; Kim et al., 2019; Lancaster et al., 2018; Liang et al., 2021).

### 3.6 MRI Measures Linked to Balance Assessments

The findings of the studies linking brain imaging with balance assessments are listed in Table 5 and are detailed in text below.

**Table 5.** Findings Reported in Studies.

| Study | Comparison | Findings |
| --- | --- | --- |
| Adam et al., 2015 | BESS correlation with FA | None |
| Bittencourt et al., 2022 | Correlations between: i) findings from group differences and HISC sub-domains; ii) findings from group × HISC interaction and HISC sub-domains | **Main effect association:**<br>Functional connectivity ↑ vestibular complaints at left MTG in Cognitive/language network<br>**Interaction association:**<br>**Withing mTBI group:** Functional connectivity ↑ vestibular complaints at anterior frontal gyrus & middle occipital gyrus in visual/cerebellar network<br>**Withing mTBI group:** Functional connectivity ↑ vestibular complaints at cuneus in visual/cerebellar network<br>**Withing mTBI group:** Functional connectivity ↓ vestibular complaints at cerebellum VI and crus I in visual/cerebellar network<br>**Within Healthy:** Functional connectivity ↑ vestibular complaints at cerebellum VI and crus I in visual/cerebellar network |
| Caeyenberghs et al., 2010 | FA TBI correlation with balance | **Eyes open- fixed surface & surround:**<br>FA ↓ with poor balance: SCP, ALIC, Cerebellum, Medial Lemniscus<br>**Eyes open- fixed surface, sway referenced visual surround:**<br>FA ↓ with poor balance: MCP |
| Caeyenberghs et al., 2011 | FA TBI correlation with balance | FA ↓ with poor balance: CST |
| Caeyenberghs et al., 2012 | Correlation of Connectivity Degree with balance | **Composite SOT Score:**<br>Connectivity Degree ↓ with poor balance: Superior Parietal Gyrus (SPG)<br>**Compromised visual and proprioception condition:**<br>Connectivity Degree ↓ with poor balance: SPG, Cerebellar Lobule IX |
| Caeyenberghs et al., 2015 | Correlation of Functional Connectivity Density with balance | **Eyes open & Compromised Proprioception (within TBI):**<br>Long-range FCD ↓ with poor balance in L Putamen (MNI: -15, 9, -9) |
|  |  | **Eyes open & Compromised Proprioception (All participants – controls & TBI):**<br>Long-range FCD ↓ with poor balance in L Putamen (MNI: -15, 9, -9)<br>**Compromised visual and proprioception condition (within TBI):**<br>Long-range FCD ↓ with poor balance in R Cerebellar Vermis I-II (MNI: 3, -36, -15)<br>**Compromised visual and proprioception condition (All participants – controls & TBI):**<br>Long-range FCD ↓ with poor balance in R Cerebellar Vermis I-II (MNI: 3, -36, -15) & R Cerebellum III (MNI: 15, -33, -21) |
| Calzolari et al., 2021 | Area 95% Ellipse during standing over soft surface with eyes-closed linked to diffusion measures | **FA correlation with balance in TBI:**<br>FA ↓ with poor balance: GCC<br>**Right:** CP, PLIC, RLIC, ACR, PCR, PTR, SS, EC, SLF, TAP<br>**Left:** ACR<br>**MD correlation with balance in TBI:**<br>MD ↑ with poor balance: SCC<br>**Right:** RLIC, SCR, PCR, PTR, SS, EC, fornix<sup>a</sup>, SLF<br>**Left:** PCR, SLF<br>**FA Impaired Balance TBI < Preserved Balance TBI:**<br>FA ↓ with poor balance: GCC<br>**Left:** ACR<br>**MD Impaired Balance TBI > Preserved Balance TBI:**<br>MD ↑ with poor balance: GCC<br>**Left:** ACR, EC<br>**FA correlation with balance (all participants – TBI and Controls):**<br>FA ↓ with poor balance: GCC, BCC, SCC, fornix<sup>b</sup><br>**Right:** CP, ALIC, PLIC, RLIC, ACR, SCR, PCR, PTR, SS, EC, Cingulum, fornix<sup>a</sup>, SLF, SFOF, UNC, TAP<br>**Left:** CP, PLIC, RLIC, ACR, SCR, PCR, PTR, SS, EC, fornix<sup>a</sup>, SLF, TAP |
|  |  | **MD correlation with balance (all participants – TBI and Controls):**<br>MD ↑ with poor balance: GCC, BCC, SCC<br>**Right:** ALIC, RLIC, ACR, SCR, PCR, PTR, SS, EC, fornix<sup>a</sup>, SLF, SFOF, UNC, TAP<br>**Left:** ALIC, RLIC, ACR, SCR, PCR, PTR, SS, EC, fornix<sup>a</sup>, SLF, TAP<br>**FA impaired balance TBI < Controls:**<br>FA ↓ with poor balance: GCC, BCC, SCC, fornix<sup>b</sup><br>**Right:** CP, PLIC, RLIC, ACR, SCR, PCR, PTR, SS, EC, Cingulum, fornix<sup>a</sup>, SLF, SFOF, UNC, TAP<br>**Left:** ALIC, RLIC, ACR, SCR, PCR, PTR, SS, EC, fornix<sup>a</sup>, SLF, UNC, TAP<br>**MD impaired balance TBI > Controls:**<br>FA ↓ with poor balance: GCC, BCC, SCC, fornix<sup>b</sup><br>**Right:** RLIC, ACR, SCR, PCR, PTR, SS, EC, Cingulum, fornix<sup>a</sup>, SLF, UNC, TAP<br>**Left:** RLIC, ACR, SCR, PCR, PTR, SS, EC, Cingulum, fornix<sup>a</sup>, SLF, UNC |
| Delano-Wood et al., 2015 | NSI (vestibular subscale) with FA value | FA ↓ with poor vestibular sub-scale score in Pontine Tegmentum |
| Diez et al., 2017 | Composite Score of SOT with dynamic BOLD activation | Prefrontal network |
| Drijkoningen et al., 2015a | SOT, LOS, RWS with FA and MD | **Baseline TBI (before training):**<br>FA ↓ with poor balance (RWS): Cerebellum and SCP<br>MD ↑ with poor balance (RWS): MCP and SCP<br>**Baseline FA link with improved balance (LOS) after training:**<br>Baseline FA ↓ in ICP<br>**Increased FA with improved balance (RWS):**<br>ICP |
| Drijkoningen et al., 2015b |  | **RWS in TBI (Grey matter):**<br>Grey matter volume ↓ with poor balance in vermal and paravermal regions of lobules I-IV, V, and VI. |
|  |  | Cerebellum Lobule I-IV (MNI: -1, -55, -17; 9, -52, -20)<br>Cerebellum Lobule VI (MNI: -7, -65, -17; -7, -72, -22)<br>Cerebellum Lobule V (MNI: -8, -60, -10; -11, -58, -23)<br>**RWS in all Participants (Grey Matter):**<br>Grey matter volume ↓ with poor balance in vermal and paravermal regions of lobules I-IV, V, and VI.<br>Cerebellum Lobule I-IV (MNI: -1, -55, -18; 8, -53, -20; -8, -66, -17; -6, -52, -22)<br>Cerebellum Lobule VI (MNI: -17, -69, -16)<br>Cerebellum Lobule V (MNI: -7, -57, -17; -11, -59, -13; -12, -58, -22)<br>**RWS in all Participants (White matter):**<br>White matter volume ↓ with poor balance in pons, dorsal medulla, mid-brain (tegmentum), SCP, MCP/Pons, MCP/Cerebellar white matter.<br>MCP/Cerebellar white-matter (MNI: -28, -49, -33; 30, -46, -34)<br>MCP/Pons (MNI: -11, -39, -31; 2, -19, -37)<br>SCP (MNI: 6, -37, -24; 11, -36, -23)<br>Pons (MNI: -2, -23, -34)<br>Medulla (MNI: 6, -44, -42)<br>**SOT in all Participants (White matter):**<br>White matter volume ↓ with poor balance in pons and MCP.<br>MCP/Pons (MNI: 2, -19, -38; -9, -33, -39; 1, -28, -38; -9, -38, -32; 3, -33, -28; 5, -37, -25; -13, -38, -42) |
| Handiru et al., 2021 | FA, MD, MA correlated with BBS & COP | None |
| Hou et al., 2022 | Change in RSFC linked with change in SOT score | None |
| Kaushal et al., 2019 | Global connectivity (Average nodal strength) correlated with BESS | None |
| Kim et al., 2019 | Axial diffusivity with balance score during standing on soft surface with eyes closed | AD ↓ in left ICP |
| Lancaster et al., 2018 | DTI measures (MD or AD) correlated with BESS | None |
| Liang et al., 2021 | Fixel based metrics correlated with SOT score | None |
| Wang et al., 2019 | Relative cerebral blood flow with BESS | rCBF ↑ with poor balance in left occipital gyrus (MNI: -34.1, -87.5, 17.8) |

None: no statistically significant differences; BESS: balance error scoring system; SOT: sensory organization test; LOS: limits of stability; RWS: rhythmic weight shifting; NSI: neurobehavioral symptom inventory; BBS: Berg balance scale; FA: fractional anisotropy; FCD: functional connectivity density; MD: mean diffusivity; AD: axial diffusivity; ALIC: anterior limb of internal capsule; CST: corticospinal tract; GCC: genu of corpus callosum; BCC: body of corpus callosum; SCC: splenium of corpus callosum; CP: cerebral peduncles; PLIC: posterior limb of internal capsule; RLIC: retrolenticular internal capsule; ACR: anterior corona radiata; PCR: posterior corona radiata; PTR: posterior thalamic radiations; SS: sagittal stratum; EC: external capsule; SLF: superior longitudinal fasciculus; SFOF: superior fronto-occipital fasciculus; TAP: tapetum; UNC: uncinate fasciculus; SCP: superior cerebellar peduncle; MCP: middle cerebellar peduncle; ICP: inferior cerebellar peduncle; rCBF: relative cerebral blood flow; fornix<sup>a</sup>: fornix (column, body); fornix<sup>b</sup>: fornix (cres)/stria terminalis; ↑: increases/positively correlated; ↓: decreases/negatively correlated.

#### 3.6.1 Findings in Studies Using Selected ROIs

Five studies using selected ROIs (Caeyenberghs et al., 2010, 2011; Delano-Wood et al., 2015; Drijkoningen et al., 2015a; Drijkoningen et al., 2015b) reported significant link of imaging measures with balance whereas one study found no link (Hou et al., 2022). Caeyenberghs et al., 2010 reported lower fractional anisotropy (FA) values linked to poor balance of TBI patients in: 1) SCP, ALIC, cerebellum, and ML during the eyes-open with fixed surface and surround condition; 2) MCP during eyes open with fixed surface but sway referenced visual surround. Caeyenberghs et al., 2011 also found that lower FA values linked to poor balance but in the CST. Drijkoningen et al., 2015a studied the effect of balance training and found: 1) at baseline, low FA in cerebellum and SCP, and high medial diffusivity (MD) values in MCP and SCP, were linked to poor balance of TBI patients in RWS test; 2) within TBI patients, lower FA in ICP at baseline linked to improved balance scores post-training in the LOS test (uncorrected for multiple comparisons); 3) within TBI patients, the increased performance in RWS test after training was linked to increase in FA values of ICP.

Drijkoningen et al., 2015b only used cerebellum and brain stem as selected ROIs and found: 1) within TBI, worse RWS test performance was linked to lower grey matter volume in vermal and paravermal regions of cerebellum lobule I-IV, V, and VI; 2) correlation between RWS performance of all participants (controls and TBI) showed that worse RWS performance linked to lower grey matter volume in vermal and paravermal regions of cerebellum lobules I-IV, V, and VI and to lower white-matter volume in pons, dorsal medulla, midbrain (tegmentum), SCP, MCP/pons, MCP/cerebellar white-matter; 3) correlation between SOT performance of all participants (controls and TBI) showed that worse SOT performance linked to lower white matter volume in MCP/pons.

Delano-Wood et al., 2015 reported that lower FA in pontine tegmentum linked with poor vestibular sub-scale score within TBI patients, however, there were no group differences between TBI and controls in the diffusion parameters of the three selected ROIs.

#### 3.6.2 Findings in Studies Using Whole-Brain Analysis

Five studies that performed whole-brain analysis (at voxel- or ROI-level) reported no link of imaging measures with balance measures (Adam et al., 2015; Handiru et al., 2021; Kaushal et al., 2019; Lancaster et al., 2018; Liang et al., 2021). Bittencourt et al., 2022 reported link of vestibular sub-scores in HISC-scale with brain regions i.e., left mid temporal gyrus within cognitive/language resting state functional network (nb: language network contains regions considered important for vestibular processing i.e., superior, middle, and inferior temporal gyri), and cerebellum VI and crus I within visual & cerebellar resting state functional network. Caeyenberghs et al., 2012 performed a graph theoretical analysis and reported that lower connectivity degree values linked with poor balance of TBI patients in SOT test (composite score of all balance conditions) in the superior parietal gyrus; whereas in condition with compromised visual and proprioceptive feedback, low connectivity degree was linked to poor balance in superior parietal gyrus and cerebellar lobule IX. Caeyenberghs et al., 2015 reported that: 1) in condition with eyes open and compromised proprioception, poor balance scores were linked to lower long-range functional connectivity density (FCD) of left putamen within TBI patients, as well as in all participants (controls and TBI); 2) in condition with both vision and proprioception compromised, poor balance scores were linked to lower long-range functional connectivity density (FCD) of right cerebellar vermis I-II within TBI patients, right cerebllar vermis I-II, and right cerebllum III in all participants (controls and TBI).

Calzolari et al., 2021, compared acute TBI patients with impaired balance versus acute TBI patients with normal balance, whereby the normal range was based upon a healthy matched controls’ posturography performance standing on a soft-surface with eyes closed. Calzolari et al., 2021 reported that: 1) FA was lower in genu of corpus callosum and left anterior corona radiata when comparing TBI with impaired balance to TBI with normal balance; 2) MD was higher in the genu of the corpus callosum, left anterior corona radiata, and external capsule when comparing TBI with impaired balance to TBI with normal balance. The remaining findings from Calzolari et al., 2021 are detailed in Table 5 due to numerous other comparisons.

Diez et al., 2017 reported that increased dynamic BOLD activation of a prefrontal network was linked to poor balance. Kim et al., 2019 reported that poor balance score (during standing on soft-surface with eyes-closed) linked with lower axial diffusivity (AD) in left ICP. Wang et al., 2019 reported that poor scores in BESS linked with increased relative cerebral blood flow (rCBF) in left occipital gyrus in TBI.

### 3.7 Statistical considerations

All studies’ statistical analyses were reviewed and only notable practices, such as the absence of multiple comparison correction or absence of direct comparison with appropriate control groups, are mentioned here.

Caeyenberghs 2015 et al., performed correlations between the balance measures and the brain regions that were significantly different between controls and TBI patients. Notably, these correlations were performed within groups i.e., a correlation of balance measure with fucntional imaging connectivity measure within the patient group, and within the control group. Similar correlation was also performed with all participants included in same analysis (patients and controls combined). However, a direct statistical comparison of correlations, indicating whether the correlation of balance measure with imaging connectivity measure was statistically different between control and patient groups, was not listed.

From Calzolari et al., 2021, we included results from 8 (of 19) statistical comparisons reported in the study, which were relevant to this systematic review i.e., comparisons linking brain imaging measures and balance measures. Four of these comparisons were performed using FA and four using MD diffusion imaging measures. The 8 comparisons included two correlation contrasts within all TBI patients, 2 correlation contrasts including all participants (patients & controls together), two comparisons of “TBI patients with impaired balance” with healthy controls, and two comparisons within TBI patients (“TBI with impaired balance” vs “TBI with normal balance”). However, the study does not indicate that if the comparisons were corrected for the number of total comparisons performed.

Delano-wood et al., 2015 found no significant difference between TBI and controls in the functional connectivity of selected ROIs. Despite the lack of a group difference, a within patient correlation of the NSI (vestibular subscale) with FA values was performed in the study. A direct statistical comparison, indicating whether the correlation of imaging connectivity measure and balance measure in patients was statistically different from that of healthy controls, was not available.

Diez et al., 2017 performed correlation of three balance measures with connectivity of a functional network. However, the study did not explicitly state that the p-values were corrected for three comparisons.

Two of the five significant findings reported in Drijkoningen et al., 2015a (and included in this systematic review) were labeled as exploratory analysis without correction for multiple comparisons. Notably, correlations of DTI measures and balance measuers were performed separately in TBI, and the control groups, and the direct statistical comparison indicating a statistical difference in correlations of the two groups was not available.

Drijkoningen et al., 2015b linked the three balance measures (SOT, LOS, RWS) with subcortical ROIs in all participants’ grey- and white-matter as well as within TBI group only. The findings were not significant for the correlations within controls and were not reported. The study does not indicate if the p-values used in the study for indicating statistical significance, were corrected for the total number of comparisons.

Kim et al., 2019 performed a comparison of diffusion imaging scans between patient and a control group, resulting in 6 significant findings (of which 4 were unique brain regions); and then the diffusion parameters from these significant brain regions were correlated with balance scores. However, only one correlation of a DTI measure with the balance measure is reported (within TBI group) without explicit mention of correction for multiple comparisons.

Wang et al., 2019 also performed within group (patients only) correlational analysis and used a cluster height threshold of p < 0.05; but also reported that the findings did not survive correction at cluster height threshold of p < 0.01.

### 3.8 EEG & fNIRS Studies

Notably, there were 5 EEG and 3 fNIRS studies that we found as part of the systematic review (Handiru et al., 2021; Helmich et al., 2020; Helmich et al., 2016; Jacob et al., 2022; Slobounov et al., 2012; Teel et al., 2014; Thompson et al., 2005; Urban et al., 2020). The comparisons within these studies were often between TBI patients vs healthy controls (Handiru et al., 2021; Helmich et al., 2016; Teel et al., 2014; Thompson et al., 2005; Urban et al., 2020) or in patients with and without concussion (Helmich et al., 2016, 2020). EEG studies reported link of imbalance with decrease of beta, delta, and theta (Teel et al., 2014) as well as decrease of alpha (Slobounov et al., 2012). Similar decrease in delta was also reported in one other study in concussed individuals (Thompson et al., 2005). One study reported a negative correlation between theta band modularity (a graph-theoretical measure) and BBS (Handiru et al., 2021), whereas one study found no EEG changes linked to imbalance (Jacob et al., 2022). The findings of the three fNIRS (Helmich et al., 2020; Helmich et al., 2016; Urban et al., 2020) studies were predominantly in the frontal regions, mainly because the electrodes were only placed on frontal ROIs. Thus, due to limited studies, all of which focused on concussed athletes (except one), we avoided discussing these studies in context of the imaging findings in mild-moderate-severe TBI. The study characteristics and the findings from these studies are listed in Table 6 and Table 7.

**Table 6.** Study and participant characteristics (EEG & fNIRS studies)

| Study | Study Type | Patients |  |  | Controls |  |  | Mode Of Injury | Injury Severity | Injury to Testing | Injury to Scan |
| --- | --- | --- | --- | --- | --- | --- | --- | --- | --- | --- | --- |
| | | Cohort | Sample Size (Gender) | Age (Mean ± SD) | Cohort | Sample Size (Gender) | Age | | | | |
| Handiru et al., 2021 | Chronic Cross-Sectional | General Population (18-65 Y) | 17 (4 F) | Mean ± SD: 48.7 ± 12.5 Y | Healthy Controls | 15 (2 F) | Mean ± SD (n = 15): 48.7 ± 12.5 Y | NA | Mild, Moderate, Moderate-Severe, Severe | NA | NA |
| Helmich et al., 2016 | Chronic Cross-Sectional | Athletes | 20 (10 F) | 27 ± 10.4 Y | Athletes | 10 (6 F) | Mean = 26.7 ± 7.9 Y | Sport Injury | Concussion | Symptomatic group: 21 ± 21 M<br>Asymptomatic group: 28 ± 16 M | NA |
| Helmich et al., 2020 | Chronic Cross-Sectional | Athletes | 62 (22 F)<br>Asymptomatic : 31 of 62<br>Symptomatic: 31 of 62 | 27 ± 10.4 Y | NA | NA | NA | Sport Injury | Concussion | Symptomatic group: 51.1 ± 56 M<br>Asymptomatic group: 27.9 ± 47.5 M | NA |
| Jacob et al., 2022 | Chronic Cross-Sectional | Athletes | 26 Females | 30.5 ± 6.9 Y | Athletes | 28 Females | 29.8 ± 8.2 Y | Sport Injury | Concussion | NA | NA |
| Slobounov et al., 2012 | Prospective | Athletes | 49 (18 F) | NA (Range: 18-25) | NA | NA | NA | Sport Injury | Concussion | 7 days | NA |
| Teel et al., 2014 | Acute Cross-sectional | NA | 7 (5 F) | 21 ± 1 Y | NA | 12 (2 F) | 21 ± 1 Y | NA | Concussion | 8 days (5 ± 1 days) | NA |
| Thompson et al., 2005 | Chronic Cross-Sectional | Athletes | 12 Males | NA (Range: 18-25) | Athletes | 12 Males | NA (Range: 18-25) | Sport Injury | Concussion | Mean: 89.4 days (Range: 70-131 days) | NA |
| Urban et al., 2021 | Acute Cross-Sectional | Athletes | 5 (4 F) | 15.8 ± 1.3 Y | Athletes | 14 (10 F) | 15.8 ± 1.7 Y | Sport Injury | Concussion | Within 2 weeks | NA |

SD: standard deviation; F: females; Y: years; M: months; NA: not available.

**Table 7.**
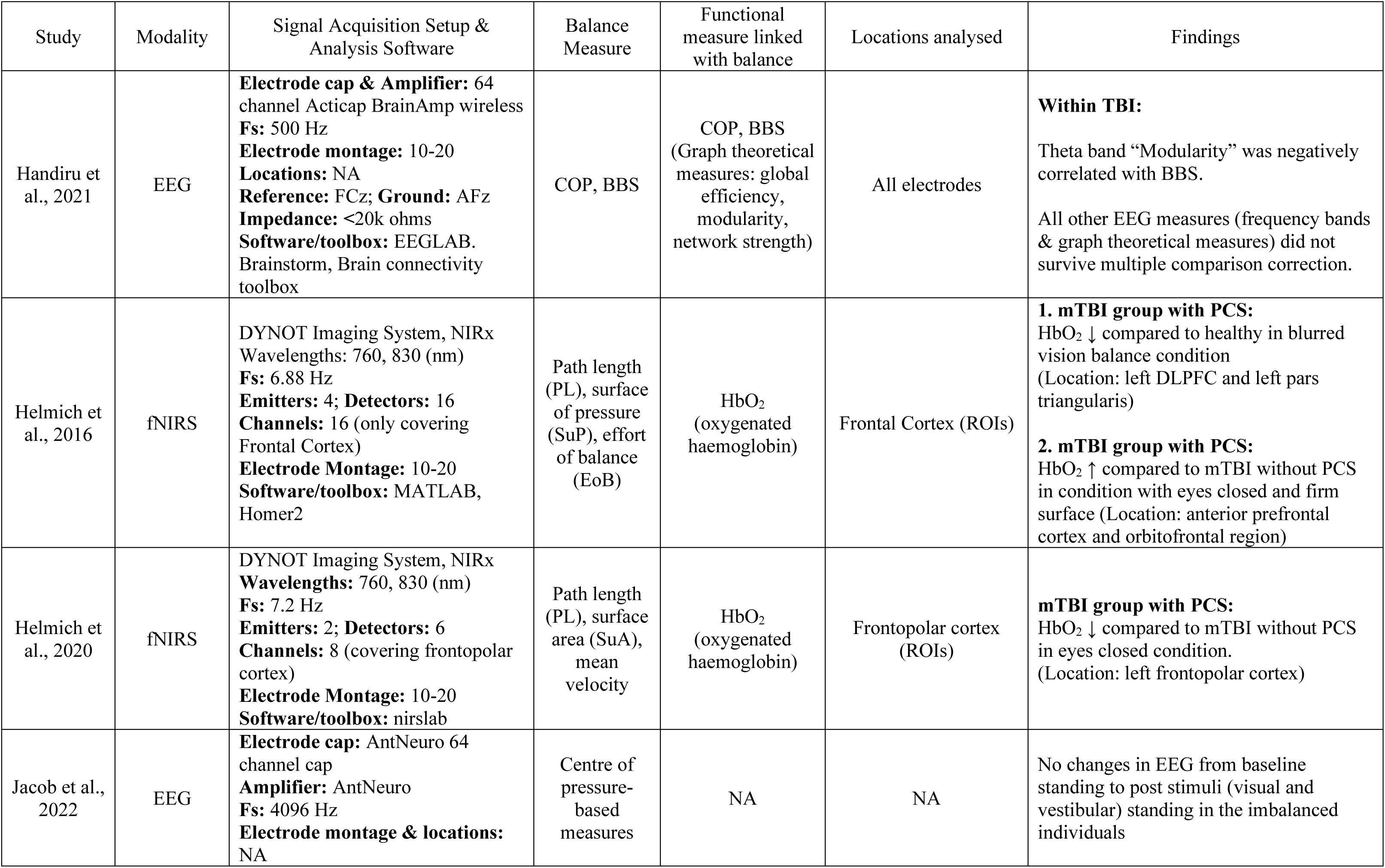
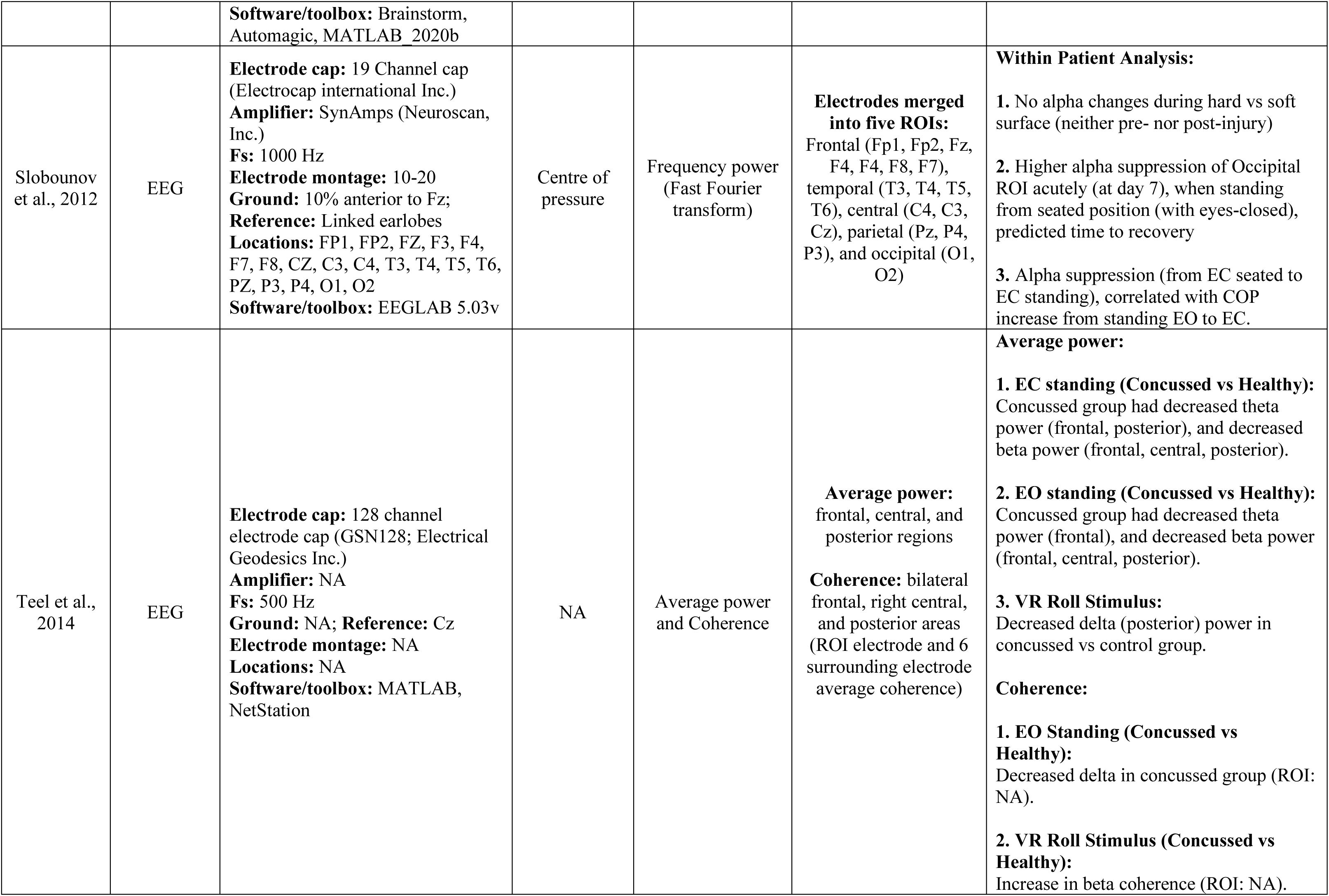
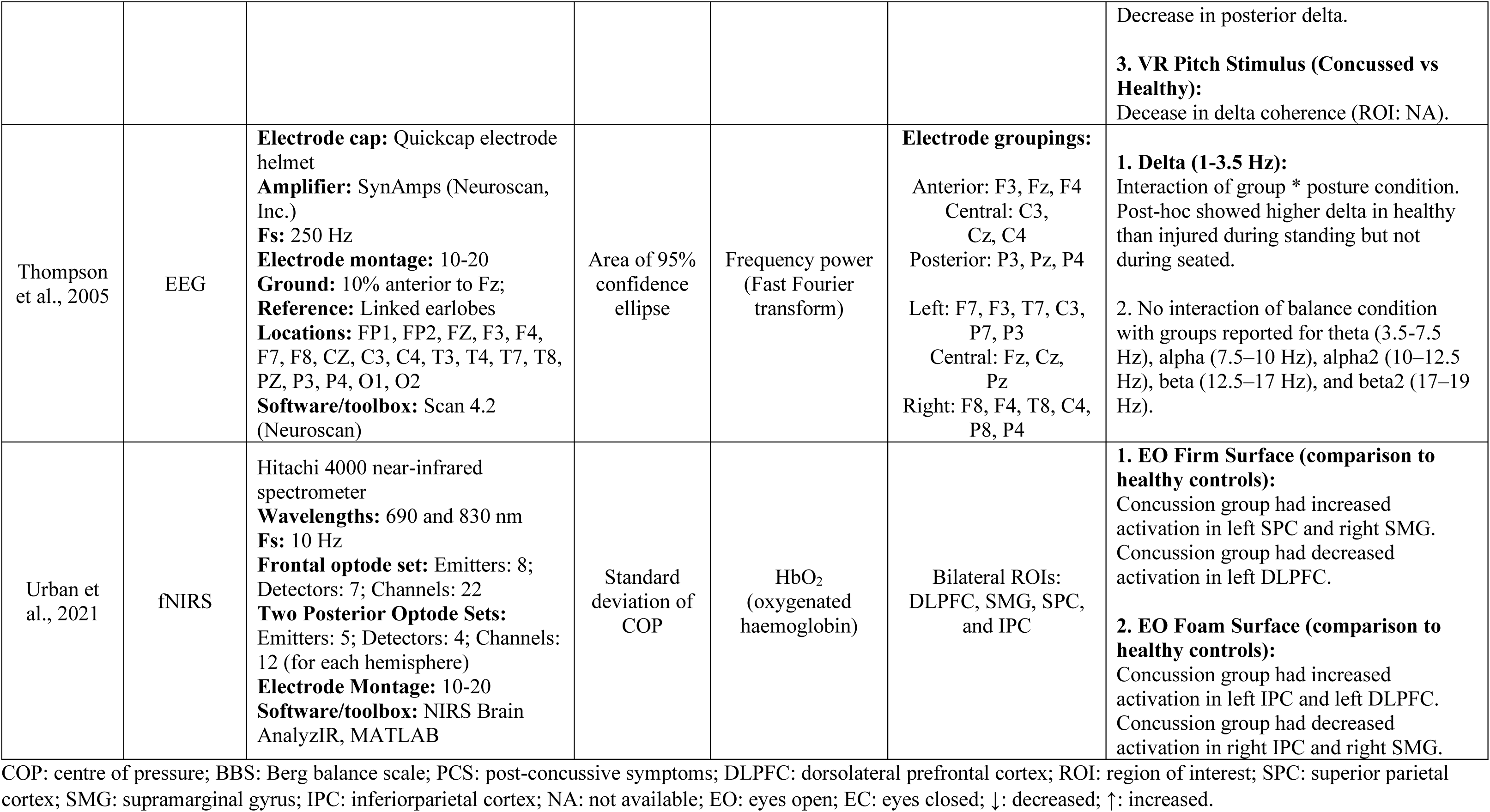
Methodology and findings (EEG & fNIRS studies)

### 3.9 Excluded Studies

There were 26 full length articles (out of 35 excluded articles) relevant to the topic that were retrieved but excluded after detailed screening as they did not fulfil inclusion criteria. They are briefly mentioned here as per the PRISMA protocol. Some studies performed a balance assessment and brain imaging but were excluded as they did not state whether an analysis linking brain imaging and balance measure is performed (Churchill et al., 2017; Churchill et al., 2021; Hammeke et al., 2013; Jang, Kim, et al., 2016; Jang, Yi, et al., 2016; Meier et al., 2020; Muftuler et al., 2020; Vartanian et al., 2021). Some of the studies either did not report data from postural balance assessment (Charney et al., 2020; Madaan et al., 2021; Madhavan et al., 2019; Sours et al., 2015; Toth et al., 2021; Weng et al., 2017) or from brain imaging (Shetty et al., 2018). One study focused on acquired brain injury and thus also had stroke patients (n = 22) combined with TBI (n = 9) (Joubran et al., 2022) whereas one study looked at working memory of TBI with impaired balance (Woytowicz et al., 2018). One study mainly looked at the link between poor sleep and number and volume of perivascular spaces (PVS) but also reported link of PVS with balance (using a symptom scale); however, the study did not report or link it with grey- or white-matter regions or networks in the brain (Piantino et al., 2021). There were 8 EEG based studies that did not fulfil inclusion criteria (Allexandre et al., 2019; Devilbiss et al., 2019; Handiru et al., 2022; Howell et al., 2018; Seo et al., 2011; Slobounov et al., 2005; van der Veen et al., 2023; Walter et al., 2017).

## 4 Discussion

This systematic review focused on identifying brain regions that are linked to postural imbalance in TBI patients, with the aim to understand underlying brain mechanisms that could explain imbalance in TBI. However, the findings from this systematic review are quite heterogeneous with low consistency in findings reported by the studies included in review. Below we first critically appraise the important methodological characteristics of studies and highlight the need for clinically well-characterized patient cohorts. We then discuss the findings reported by the studies in light of previous literature, the reproducible findings among these studies, as well as current limitations, and the recommendations for future studies.

### 4.1 Cohort – timing of recruitment, clinical assessment, and treatment

Traumatic brain injury (TBI) is the commonest cause of injury related death and disability, where c. 60 million people have a TBI every year (Maas et al., 2017, 2022) with further increased falls risk post-TBI (Elser et al., 2023). Despite the post-TBI falls related risks, we only found one acute-prospective study in moderate-severe TBI that assessed post-TBI vestibular-mediated balance problems and its brain mechanisms via neuroimaging (Calzolari et al., 2021). Given that TBI is common in relatively young adults (Calzolari et al., 2021; Marcus et al., 2019; Rust et al., 2022) and results in socio-economic costs of up to US$400 billion (Maas et al., 2022), it is also surprising to notice the lack of studies that assessed vestibular-mediated balance problems and their brain mechanisms in the general population ranging between 18-65 years (Calzolari et al., 2021; Handiru et al., 2021).

Notably, none of the studies included in this systematic review, except (Calzolari et al., 2021), performed or reported about acutely performed clinical vestibular assessment of peripheral vestibular damage or diagnoses such as benign paroxysmal positional vertigo (BPPV). Recent work shows that there is a high burden of vestibular disorders in TBI, both from central injury and peripheral injury including BPPV (Calzolari et al., 2021; Harrell et al., 2023; Marcus et al., 2019) and vestibular nerve transection (Arshad et al., 2017; Marcus et al., 2019) as well as associated diagnoses such as vestibular migraine (Marcus et al., 2019), whose commonest manifestation is objective imbalance (Von Brevern et al., 2005).

Even though the cross-sectional studies of moderate-severe TBI in this review had average time of circa three years from injury to testing, some patients were also tested as early as four months after the injury (Diez et al., 2017; Drijkoningen et al., 2015a; Drijkoningen et al., 2015b). Notably, previously reported recurrence rate of BPPV was up to 67% in TBI (Calzolari et al., 2021; Gordon et al., 2004), which was supplemented by recent studies showing that recurrence can occur in a period ranging from up to four (Smith et al., 2020) to six months (Hadi et al., 2023 – in preparation; recurrence rate of 31.6%) post-TBI. It follows that any study aiming to understand the brain correlates of imbalance should assess all these clinical features both acutely and at follow-up to exclude or treat patients for these additional vestibular diagnoses, prior to balance and imaging assessment. Additionally, some maladaptive syndromes that are linked to imbalance e.g., persistent postural perceptual dizziness or PPPD (Seemungal & Passamonti, 2018), may also develop chronically and must be controlled for in chronic TBI studies.

Notably, characterization of injury severity is key for TBI studies. In this review, multiple studies were rated as high risk on selection of participants due to lack of details on how the injury severity was defined. Combining brain imaging of mild and moderate TBI within same analysis can be problematic as moderate TBI often have more structural imaging deficits compared to mild. In that case, brain imaging findings are more likely to be driven by severe injury in moderate TBI and are less likely to reflect the brain imaging changes in mild TBI.

### 4.2 Reproducible Findings

The most consistent findings reported in this systematic review are mainly from six studies (Caeyenberghs et al., 2010, 2011, 2012, 2015; Drijkoningen et al., 2015a; Drijkoningen et al., 2015b) from the same research centre or authors, with partially overlapping patient cohorts across the studies (nb: some studies also showed a complete overlap of sample (Caeyenberghs et al., 2010, 2011, 2012) but since authors were not contacted to confirm this nor the authors declared it in the respective articles, we do not definitely state this). Caeyenberghs et al., 2010 used selected ROIs for analysis and Caeyenberghs et al., 2012 performed a whole brain analysis of DTI imaging. The cerebellum was shown to link with imbalance in both studies (Caeyenberghs et al., 2010, 2012) but findings showing link of SCP, ALIC, ML, and MCP with balance, were not reproduced in the whole brain analysis in Caeyenberghs et al., 2012. Similarly, the findings from Caeyenberghs et al., 2011 were not consistent with those reported in (Caeyenberghs et al., 2010, 2012), which could be due to a bigger sample of patients in Caeyenberghs et al., 2011 (n = 24) compared to n = 12 in (Caeyenberghs et al., 2010, 2012).

In the five studies with another partially overlapping patient cohort (Caeyenberghs et al., 2015; Diez et al., 2017; Drijkoningen et al., 2015a; Drijkoningen et al., 2015b; Liang et al., 2021), the sample size varied from having n = 12 (Caeyenberghs et al., 2015), n = 14 (Diez et al., 2017), n = 19 (Drijkoningen et al., 2015a), n = 18 (Drijkoningen et al., 2015b), and n = 17 (Liang et al., 2021). Three of these studies performed whole-brain analysis using DTI (Liang et al., 2021), resting-state (Caeyenberghs et al., 2015), and both, DTI and resting-state together (Diez et al., 2017).

Liang et al., 2021 found no associations between changes in white-matter and improved balance performance after training, and also reported no baseline associations between balance measures and diffusion imaging parameters. Notably, Diez et al., 2017 found a link of SOT score with a prefrontal functional networks, whereas, Caeyenberghs et al., 2015 reported a link of balance measures with regions such as putamen and cerebellum; yet all of these findings were not overlapping, despite the partial overlap in the sample of patients. Drijkoningen et al., 2015a and Drijkoningen et al., 2015b performed analysis within selected ROIs, mostly subcortical. Cerebellum, SCP, and MCP were found to be linked with imbalance in both studies, however, no sub-regions of cerebellum were reported in Drijkoningen et al., 2015a. The cerebellar lobules reported in Drijkoningen et al., 2015b are not consistent with findings of Caeyenberghs et al., 2015, which only found cerebellum lobule IX to be linked with imbalance.

In general, despite selecting participants from a patient cohort pool with partial or complete overlap in paticipants between studies, the findings do not overlap within these eight studies (Caeyenberghs et al., 2010, 2011, 2012, 2015; Diez et al., 2017; Drijkoningen et al., 2015a; Drijkoningen et al., 2015b; Liang et al., 2021). While the lack of reproducibility can be attributed to the methodological differences or low sample sizes, a general lack of reproducible findings in overlapping patients maybe due to factors beyond simple methodology, inlcuding maladaptive clinical disorders (Seemungal & Passamonti, 2018), which may develop over the time such that individual patients’ performance could vary over time.

### 4.3 Sample Size and Statistical Considerations

Multiple studies performed correlations between balance measures and the brain regions within groups (within controls and within TBI) and with all participants together (controls and TBI combined) (Caeyenberghs et al., 2015; Calzolari et al., 2021; Delano-Wood et al., 2015; Drijkoningen et al., 2015; Drijkoningen et al., 2015; Kim et al., 2019; Wang et al., 2019). However, a direct statistical comparison of the two groups’ correlation was often missing. Moreover, if studies performed multiple comparisons (multiple contrasts) or had multiple outcomes measures to assess balance, whether the p-values were corrected for multiple contrasts and/or the multiple balance measures was not explicitly stated.

It is worth highlighting that the analysis within TBI sub-groups could be useful, if performed between TBI patients with and without impaired balance, as it could control for TBI specific findings. Only one study in this review performed such analysis, using TBI group without imbalance (n = 11) as a control group and compared it with TBI with imbalance (n = 19) (Calzolari et al., 2021). Notably, two EEG studies (Helmich et al., 2016, 2020) also performed similar analysis to compare balance of mild TBI with and without post-concussive symptoms. However, notice that groups in this case were categorized on the basis of post-concussive symptoms rather than imbalance, and thus the EEG findings during a balance task would reflect the functional brain changes linked to the post-concussive symptoms and not due to imbalance.

The sample sizes of studies we included in this systematic review were considered low, given the recent recommendations of sample sizes for brain wide association studies (Marek et al., 2022; Spisak et al., 2023) . Thus, we include a cautionary note, to interpret the findings from the studies included in this systematic review in context of all available information.

### 4.4 Imbalance in TBI and proprioception

It is also worth mentioning that none of the studies systematically assessed proprioception. The participant exclusion criteria in studies often listed any “musculoskeletal condition”, but it was not always clear if this was a reference to post-injury “condition”. We also eluded any discussion about proprioception from this review as the knowledge about proprioceptive deficits and its link to imbalance in TBI is sparse; such that a recent study in concussed individuals claimed that the proprioception has never been systematically assessed in concussion (Lempke et al., 2023). Moreover, a search on “Web of Science” relating imbalance, TBI, and proprioception or somatosensory system (without including imaging search terms) only resulted in 29 articles. Within these 29 studies, no study systematically assessed proprioception prospectively and linking it with brain imaging.

Often studies use objective measures such as sensory organization test (SOT) or posturography to assess balance, in which the effect of proprioception is assessed by asking the participants to stand over a “soft surface” such as foam, and thus compromising the proprioceptive feedback. However, this measures the combined contribution of vision and proprioception; and in case where visual feedback is not available (such as when eyes are closed), the combined contribution of vestibular system and proprioception is assessed. Thus, a systematic assessment of proprioception is needed in future studies, which can shed some light on proprioceptive contribution in deficits of postural balance post-TBI.

### 4.5 Current Evidence of Neural Correlates of Imbalance in TBI

Below we discuss the findings of this systematic review in context of previous literature.

#### 4.5.1 Cerebellum

The recurring finding from all included studies was cerebellum, which was reported in three studies that performed whole-brain analysis (Bittencourt et al., 2022; Caeyenberghs et al., 2012, 2015), and in three studies that used cerebellum as an ROI (Caeyenberghs et al., 2010; Drijkoningen et al., 2015a; Drijkoningen et al., 2015b). However, among these studies, the sub-regions localised within the cerebellum were not consistent. Caeyenberghs et al., 2010 used cerebellum (as a whole) in ROI analysis and thus no specific sub-region is reported.

Caeyenberghs et al., 2012 reported lower connectivity degree (a structural connectivity measure) linked to poor balance in cerebellar lobule IX, which is considered an important site for processing vestibular signals in humans and is often termed as vestibular cerebellum (Barmack, 2003; Kheradmand & Zee, 2011; Nigmatullina et al., 2015). Skilled dancers with high balance ability are also reported to have lower grey matter volume in cerebellum lobule IX (Nigmatullina et al., 2015), which could reflect the important role of cerebellum lobule IX in vestibular function including postural balance. An important consideration is the impact of injury distant to the site of the observed brain imaging effect. In this regard, one study showed that balance impaired individuals due to incomplete spinal cord injury, had reduced volume in cerebellum lobule IX (Villiger et al., 2015), implying a component of this atrophy is secondary Wallerian degeneration from disrupted proprioceptive signal input following spinal cord injury (and hence, it further emphasizes the need for detailed clinical assessment to account for or exclude peripheral vestibular or proprioceptive dysfunction).

Caeyenberghs et al., 2015 reported involvement of cerebellar vermis I-II in mediating postural imbalance in TBI. Cerebellar vermis is considered to be the cerebellum’s most involved zone in mediating postural control (Dijkstra et al., 2020). A systematic review and meta-analysis, looking at studies that performed a mental imagery of a postural task in healthy individuals, reported that 6 out of 8 included studies reported activation in cerebellar vermis (Dijkstra et al., 2020). One study, using mobile PET scan while standing on one foot (Ouchi et al., 1999), showed an increased activation of anterior cerebellar vermis, highlighting the active role of this region in mediating the postural balance.

Drijkoningen et al., 2015a did not report a sub-region in cerebellum whereas Drijkoningen et al., 2015b found that reduced grey- and white-matter volume in cerebellar lobules I-IV, V, and VI were linked to imbalance. Another study included in this review (Bittencourt et al., 2022) reported that post-TBI vestibular symptoms (including imbalance) were linked to altered connectivity of cerebllar lobule VI and crus I. In individuals with balance impairment (incomplete spinal injury (Villiger et al., 2015) & Parkinson’s disease (Sehm et al., 2014)), balance training caused an increase in volume of crebellum lobules V and VI, suggesting their link with balance training and their possible role in post-TBI imbalance. Similarly, reduced volume in cerebellum lobules IV, V, and VI have also been linked to postural imbalance in multiple sclerosis (Prosperini et al., 2013).

#### 4.5.2 Cerebellar Peduncles

The second most consistent finding from the included studies was the cerebellar peduncles including superior, middle, and inferior cerebellar peduncles (SCP, MCP, ICP). SCP and MCP are reported to be linked with imbalance in TBI in the three studies (Caeyenberghs et al., 2010, Drijkoningen et al., 2015a, Drijkoningen et al., 2015b), all of which used SCP and MCP tracts as ROIs whereas ICP was linked with imbalance in two studies (Drijkoningen et al., 2015a, Kim et al., 2019).

One of the study reported that the FA value in ICP increased with balance training (Drijkoningen et al., 2015a). In contrast, a previous study in multiple sclerosis patients with impaired balance (Prosperini et al., 2014), reported that ICP did not show significant improvement in DTI parameters in response to balance training, whereas an increase in FA and radial diffusivity (RD) in SCP was observed. Thus, SCP and ICP seems to be linked with imbalance and may also respond to balance training whereas MCP is only reported to be linked with imbalance on baseline testing of TBI patients but not to balance training. Note that we cannot systematically suggest that MCP is not involved when performing balance training as it is implausible.

Multiple studies in MS patients have previously reported the link between postural deficits and low FA and increased RD values in SCP (Gera et al., 2020), MCP (Odom et al., 2021), and ICP (Gera et al., 2020; Odom et al., 2021); notably, both these studies also used analysis in which cerebellar peduncles were selected as ROIs. Similar association of diffusion parameters with SCP and MCP has also been reported in Parkinson’s disease patients (using SCP, MCP, and ICP as ROIs) with freezing of gait (Bharti et al., 2019), suggesting their link with postural balance in neurodegeneration.

In general, there is existing evidence of altered diffusion parameters of cerebellar peduncles and linked imbalance in neurodegeneration. Notably, however, the studies that used these tracts (SCP, MCP, ICP) as ROIs were often the ones that reported them to be linked with balance (Caeyenberghs et al., 2010; Drijkoningen et al., 2015a; Drijkoningen et al., 2015b), which is important to consider in terms of analysis to avoid selection bias in reported findings.

#### 4.5.3 Other Inconsistent Reported Findings

Most of the studies included in this systematic review reported brain regions or white-matter tracts which were not reported in any of the other MRI studies. These included medial lemniscus (Caeyenberghs et al., 2010), corticospinal tract (Caeyenberghs et al., 2011), superior parietal gyrus (Caeyenberghs et al., 2012), left putamen (Caeyenberghs et al., 2015), a pre-frontal functional network (Diez et al., 2017), pons and medulla (Drijkoningen et al., 2015b), left occipital gyrus (Wang et al., 2019), pontine tegmentum (Delano-Wood et al., 2015), and ALIC (Caeyenberghs et al., 2010; Calzolari et al., 2021).

Calzolari et al., 2021 reported four comparisons in which the diffusion parameters of ALIC were reportedly linked to imbalance. These included: i) two direct comparisons of TBI patients with healthy controls (one with FA and one with MD); and ii) two correlations of all participants’ (TBI and controls combined) diffusion parameters with balance measure (one with FA and one with MD). In contrast, when diffusion imaging parameters of TBI patients with impaired balance were compared with TBI patients without impaired balance, FA and MD values of ALIC were not different between these two groups. Thus, it seems that ALIC is damaged in TBI (as it has attenuated parameters compared to controls) but might not link with imbalance.

Calzolari et al., 2021 also compared TBI patients with and without balance impairment, which is more likely to control for TBI specific findings. This showed that TBI patients with impaired balance had low FA and high MD values in genu of corpus callosum and left anterior corona radiata. Left external capsule also showed increased MD values in TBI patients with impaired balance. Damage to corpus callosum (CC) could have a major role in causing postural imbalance in TBI since TBI often suffer damage to CC (Ghajari et al., 2017; Jolly et al., 2020). FA values in genu of CC are previously reported to be linked with deficits of dynamic balance in older adults (Rosario et al., 2016). Additionally, there is evidence to suggest that vestibular pathways are connected bihemispherically via the corpus callosum (Kirsch et al., 2016). Given the bihemispheric involvement in balance control, few would argue against the involvement of the CC in human postural control and its damage in TBI as a key mediator of imbalance after TBI.

The superior parietal gyrus, shown to link with imbalance in Caeyenberghs et al., 2012, reportedly activates in response to vestibular stimulation (Della-Justina et al., 2014; Helmchen et al., 2020), and the degree of its functional activation also links with the postural stability (Mitsutake et al., 2020). Similarly, inhibition of parietal regions using non-invasive brain stimulation (tDCS) has also been shown to modulate postural balance in healthy participants (Oka et al., 2022). Note however that inferior parietal activation is more commonly reported in response to vestibular stimulation (Bense et al., 2001; Eickhoff et al., 2006; Helmchen et al., 2020) and also shows structural changes in response to balance training (see review – Surgent et al., 2019).

Wang et al., 2019 reported a link between poor balance and relative cerebral blood flow (rCBF) in the left occipital gyrus in TBI. A similar finding, however in middle occipital gyrus, was also found by Bittecourt et al., 2022. Previous studies have reported the link between balance training and structural changes in the left inferior occipital gyrus in healthy (Taubert et al., 2010) and left middle occipital gyrus in patients (Villiger et al., 2015). Superior (Helmchen et al., 2020), middle (Aedo-Jury et al., 2020; Della-Justina et al., 2014; Helmchen et al., 2020), and inferior occipital gyri (Bense et al., 2001; Della-Justina et al., 2014; Helmchen et al., 2020) are also shown to have increased activation in response to vestibular stimulation. Whether these regions which appear to mediate multisensory integration, have a specific role in postural balance is unclear.

Caeyenberghs et al., 2015 reported a link between imbalance and the left putamen and Diez et al., 2017 reported involvement of a frontal functional network (which also included caudate). Activation of striatal regions (caudate and putamen) (Helmchen et al., 2020; Mitsutake et al., 2020) and the frontal regions (Bense et al., 2001; Della-Justina et al., 2014; Helmchen et al., 2020; Lobel et al., 1998) is also commonly reported in vestibular stimulation studies. A systematic review in healthy individuals during a postural task (Dijkstra et al., 2020) reported that one study showed functional activation in putamen (Jahn et al., 2004) and two in caudate (Karim et al., 2014; Zwergal et al., 2012) in response to mental imagery or observation of a postural task. Moreover, nearly all studies report activation in frontal regions (Dijkstra et al., 2020) during mental imagery of a postural task. One PET study has reported activation of bilateral middle frontal gyri during standing with eyes closed while scanning, suggesting possible causal role of frontal gyri in control of postural balance (Ouchi et al., 1999).

We do not discuss in detail the medial lemniscus, corticospinal tract, and midbrain regions (medulla and pons). All of these subcortical regions are important in motor function and postural balance due to their involvement in bottom-up proprioceptive signals, however, only the studies using these tracts as ROIs found their link with postural imbalance in TBI. Nonetheless, the pedunculopontine nucleus (PPN) – a region located in upper pons, is a highly vestibular responsive cholinergic nucleus (Aravamuthan & Angelaki, 2012) and could have causal role in human postural control (Yousif et al., 2016). There is evidence suggesting that cholinergic PPN neuronal loss (not dopaminergic) is linked to postural imbalance in Parkinson’s Disease (PD) (Bohnen et al., 2012; Bohnen & Albin, 2009). A recent study in a large sample of PD patients showed a link between thalamic cholinergic loss and postural imbalance but not with cortical cholinergic or dopaminergic denervation (Bohnen et al., 2022).

Notably, none of the studies except Bittencourt et al., 2022 found involvement of temporal regions. The temporal gyri are known to be of considerable importance in processing vestibular sensations as indicated by an invasive electrical stimulation study (Kahane et al., 2003). The lack of focal finding in temporal gyrus in most studies could reflect the notion of network disruption to be the cause of most vestibular problems as shown previously (Calzolari et al., 2021; Hadi et al., 2022).

## 5 Conclusions and Recommendations

Our findings indicate that the mechanisms underlying postural deficits in TBI are poorly understood. Given the multiple levels of vulnerability ranging from the vestibular organ, the nerve, brainstem and cerebellar circuits, thalamic relay areas, and cortical processing pathways, it is likely that traumatic brain injury could result in imbalance due to multiple reasons. This could also explain the inconsistency in findings of current literature. Thus, apart from removing peripheral diagnoses (vestibular organ & nerve related problems) from the equation, studies should account for multiple models of postural control to control for biased reporting or inconsistent findings. Individual studies looking at subcortical function separate from cortical function may also produce findings biased towards certain regions thus a whole-brain analysis approach is recommended.

Lack of studies in general population with TBI, and lack of studies evaluating postural imbalance in TBI is surprising, considering that 50 million people worldwide have a TBI in a year (Maas et al., 2017, 2022). Vestibular dysfunction in TBI can go unnoticed since patients’ subjective reports of their balance do not correlate well with clinical and objective assessment of balance (Marcus et al., 2019; Sargeant et al., 2018; Wood et al., 2022). Thus, assessment of vestibular dysfunction is key in TBI patients, which could be a marker of prognosis post TBI (Schlosser et al., 2009).

Finally, none of the studies included in this review accounted for the lesions and it is possible that the lesions and their spatial location, might explain the findings in the studies and could also be a predictor of imbalance. In contrast, the extensive involvement of cortical regions mediating postural control indicates significant neural redundancy hence, it seems that critical nodes whose damage would compromise balance control is less likely than the impact of network disconnection. It is also important to state that sample size in the studies were low, which could be one of the reasons of lack of replicable findings across studies with independent samples.

## Data Availability

All data produced in the present study is summarized in the form of tabulated information and is provided along with the manuscript.

## Conflict of Interest

The authors declare that they have no conflict of interest.

## Funding

The Medical Research Council (MRC), The Jon Moulton Charity Trust, The US Department of Defense - Congressionally Directed Medical Research Program (CDMRP).

## Author Contributions

**Zaeem Hadi:** Conceptualization, Investigation, Methodology, Software, Formal analysis, Visualization, Writing - Original Draft. **Mohammad Mahmud:** Methodology, Writing – review and editing. **Barry M Seemungal:** Project administration, Funding acquisition, Resources, Supervision, Writing – review and editing.

## Appendix

### Search terms for Web of Sciences (Core collection)

1. ((balance) OR (imbalance)) OR (posture*)
2. ((traumatic brain injury) OR (TBI)) OR (Concussion*)
3. ((((((((((((((((((brain imaging) OR (neuroimaging)) OR (diffusion tensor imaging)) OR (DTI)) OR (voxel based morphometry)) OR (VBM)) OR (functional imaging)) OR (structural imaging)) OR (magnetic resonance imaging)) OR (fMRI)) OR (magnetic resonance spectroscopy)) OR (single photon emission computed tomography)) OR (Positron emission tomography)) OR (Electroencephalography)) OR (EEG)) OR (Magnetoencephalography)) OR (Near-Infrared Spectroscopy)) OR (Evoked Potentials)) OR (Tomography, X-Ray Computed)
4. NIRS OR PET OR MRS OR SPECT OR CT OR fNIRS
5. #3 OR #4
6. #1 AND #2 AND #5

### Search terms for Web of Sciences (All databases: BIOSIS Citation Index, CABI: CAB Abstracts, KCI-Korean Journal Database, MEDLINE, Preprint Citation Index, SciELO Citation Index)

1. ((balance) OR (imbalance)) OR (posture*)
2. ((traumatic brain injury) OR (TBI)) OR (Concussion*)
3. ((((((((((((((((((brain imaging) OR (neuroimaging)) OR (diffusion tensor imaging)) OR (DTI)) OR (voxel based morphometry)) OR (VBM)) OR (functional imaging)) OR (structural imaging)) OR (magnetic resonance imaging)) OR (fMRI)) OR (magnetic resonance spectroscopy)) OR (single photon emission computed tomography)) OR (Positron emission tomography)) OR (Electroencephalography)) OR (EEG)) OR (Magnetoencephalography)) OR (Near-Infrared Spectroscopy)) OR (Evoked Potentials)) OR (Tomography, X-Ray Computed)
4. NIRS OR PET OR MRS OR SPECT OR CT OR fNIRS
5. #3 OR #4
6. #1 AND #2 AND #5

### Search terms for Scopus

((((balance) OR (imbalance)) OR (posture*)) AND (((traumatic AND brain AND injury) OR (tbi)) OR (concussion*))) AND (((((((((((((((((((((((((brain AND imaging) OR (neuroimaging)) OR (diffusion AND tensor AND imaging)) OR (dti)) OR (voxel AND based AND morphometry)) OR (vbm)) OR (functional AND imaging)) OR (structural AND imaging)) OR (magnetic AND resonance AND imaging)) OR (fmri)) OR (magnetic AND resonance AND spectroscopy)) OR (single AND photon AND emission AND computed AND tomography)) OR (positron AND emission AND tomography)) OR (electroencephalography)) OR (eeg)) OR (magnetoencephalography)) OR (near-infrared AND spectroscopy)) OR (evoked AND potentials)) OR (tomography, AND x-ray AND computed)) OR (pet)) OR (mrs)) OR (nirs)) OR (spect)) OR (ct)) OR (fnirs))

### Search terms for Pubmed

((((balance) OR (imbalance)) OR (posture*)) AND (((traumatic brain injury) OR (TBI)) OR (Concussion*))) AND ((NIRS OR PET OR MRS OR SPECT OR CT OR fNIRS) OR (((((((((((((((((((brain imaging) OR (neuroimaging)) OR (diffusion tensor imaging)) OR (DTI)) OR (voxel based morphometry)) OR (VBM)) OR (functional imaging)) OR (structural imaging)) OR (magnetic resonance imaging)) OR (fMRI)) OR (magnetic resonance spectroscopy)) OR (single photon emission computed tomography)) OR (Positron emission tomography)) OR (Electroencephalography)) OR (EEG)) OR (Magnetoencephalography)) OR (Near-Infrared Spectroscopy)) OR (Evoked Potentials)) OR (Tomography, X-Ray Computed)))

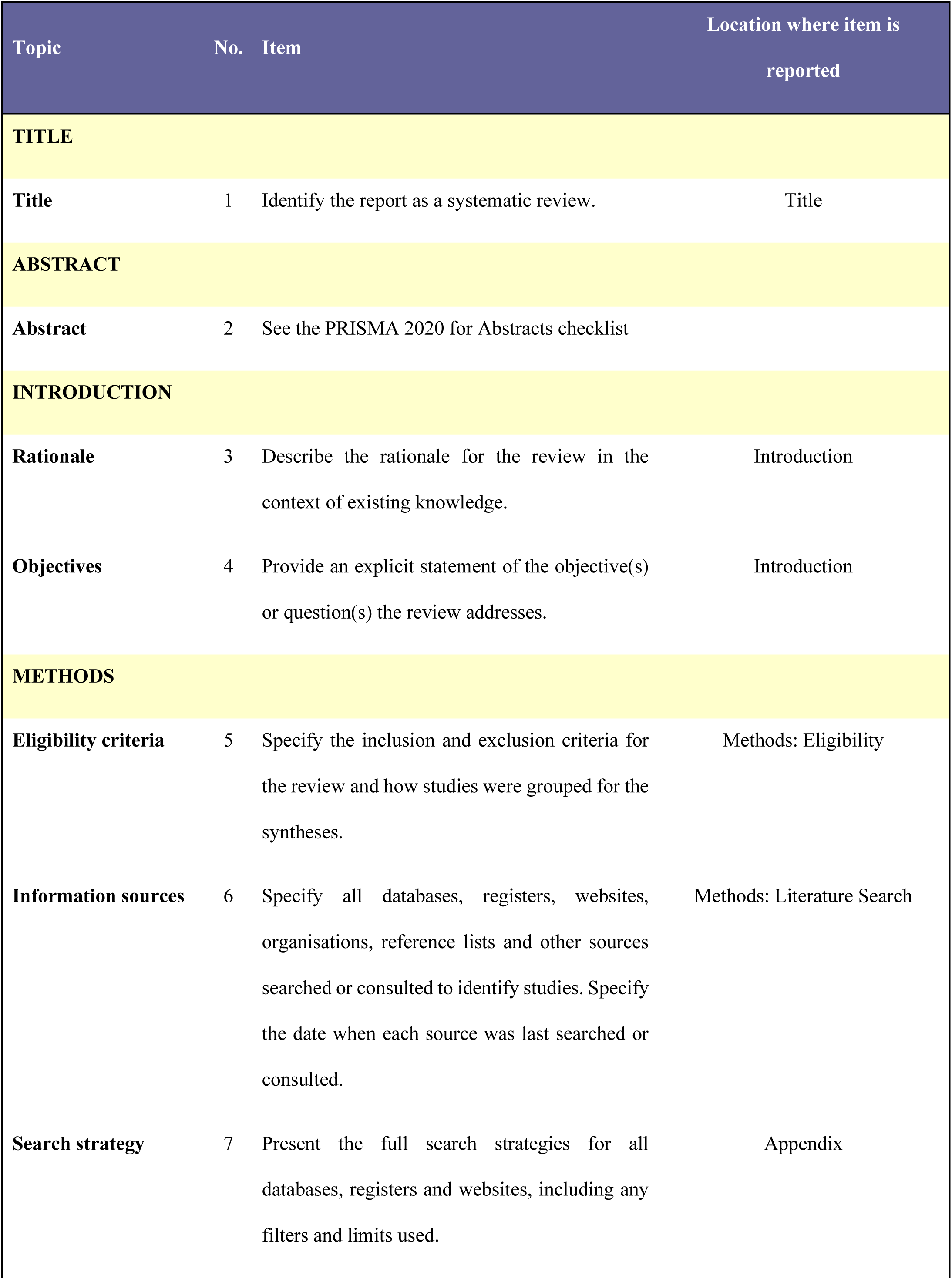
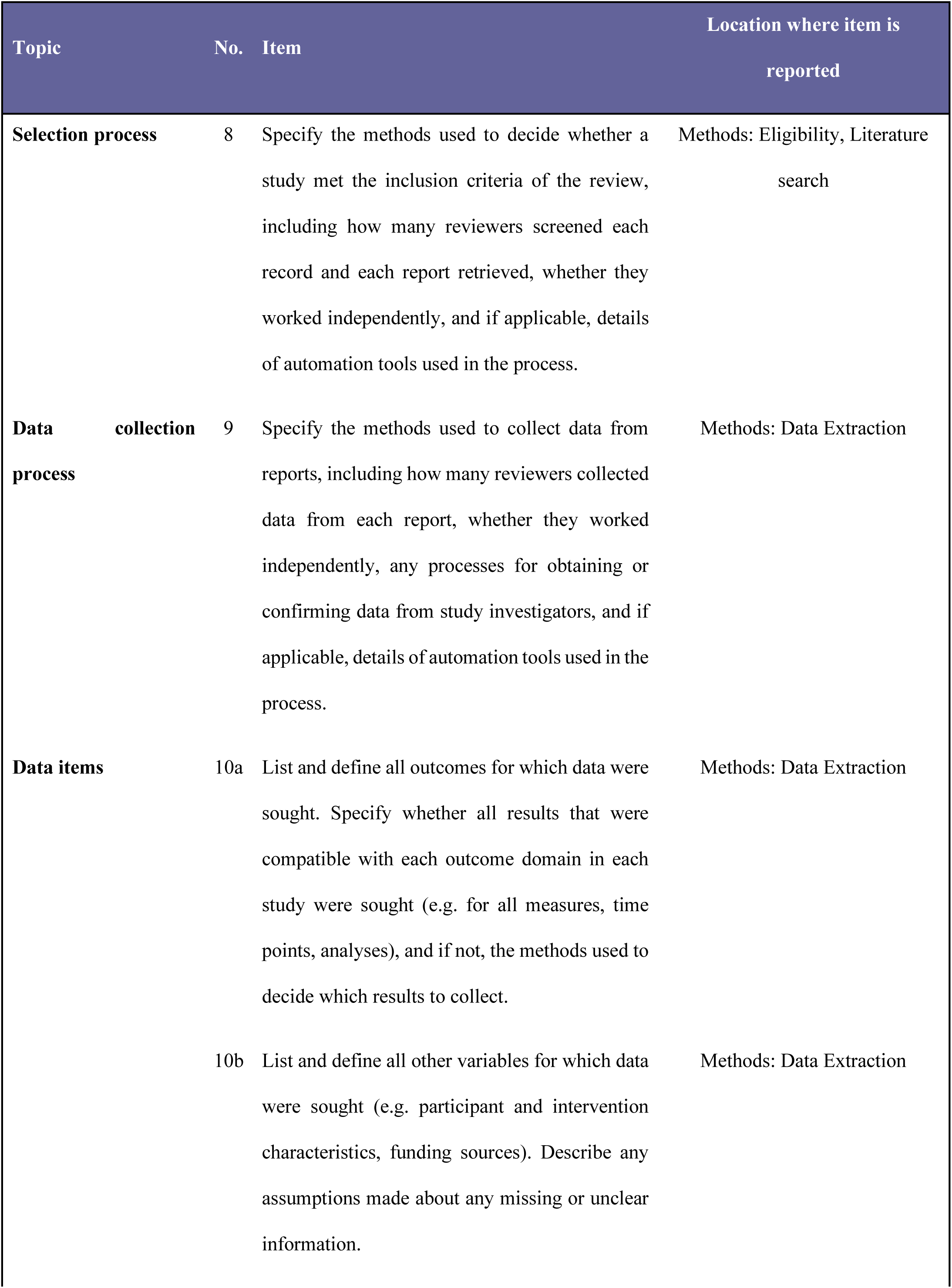
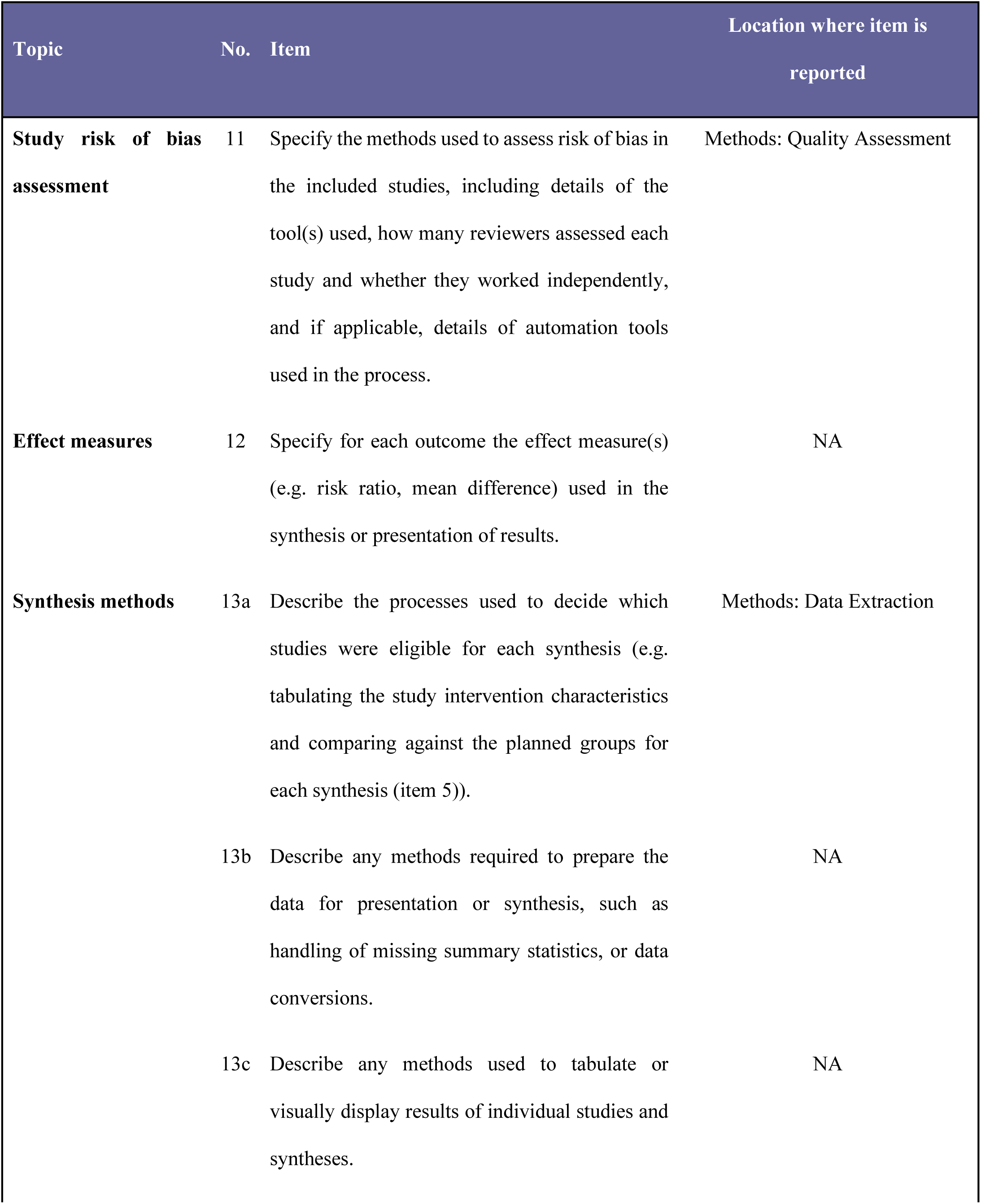
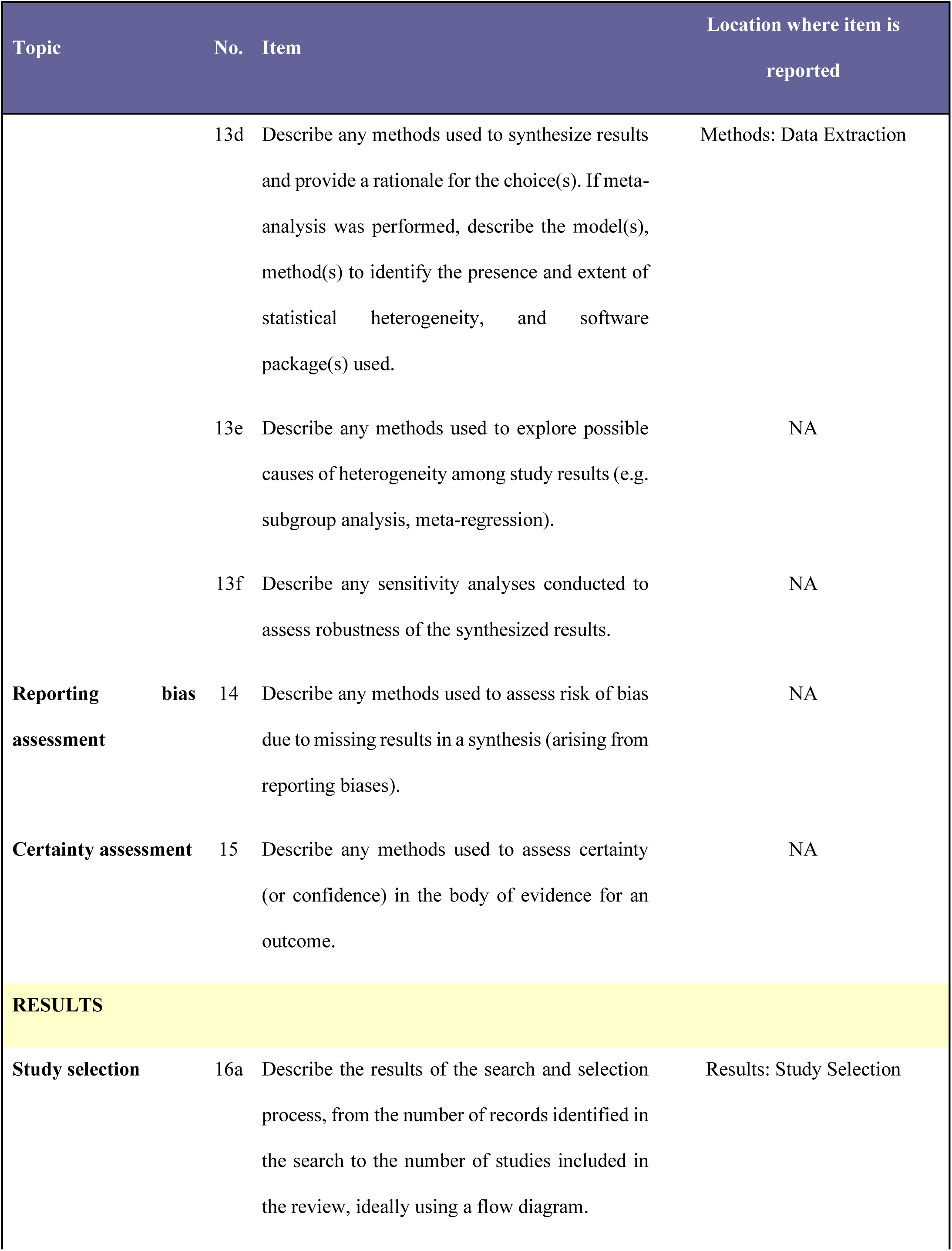
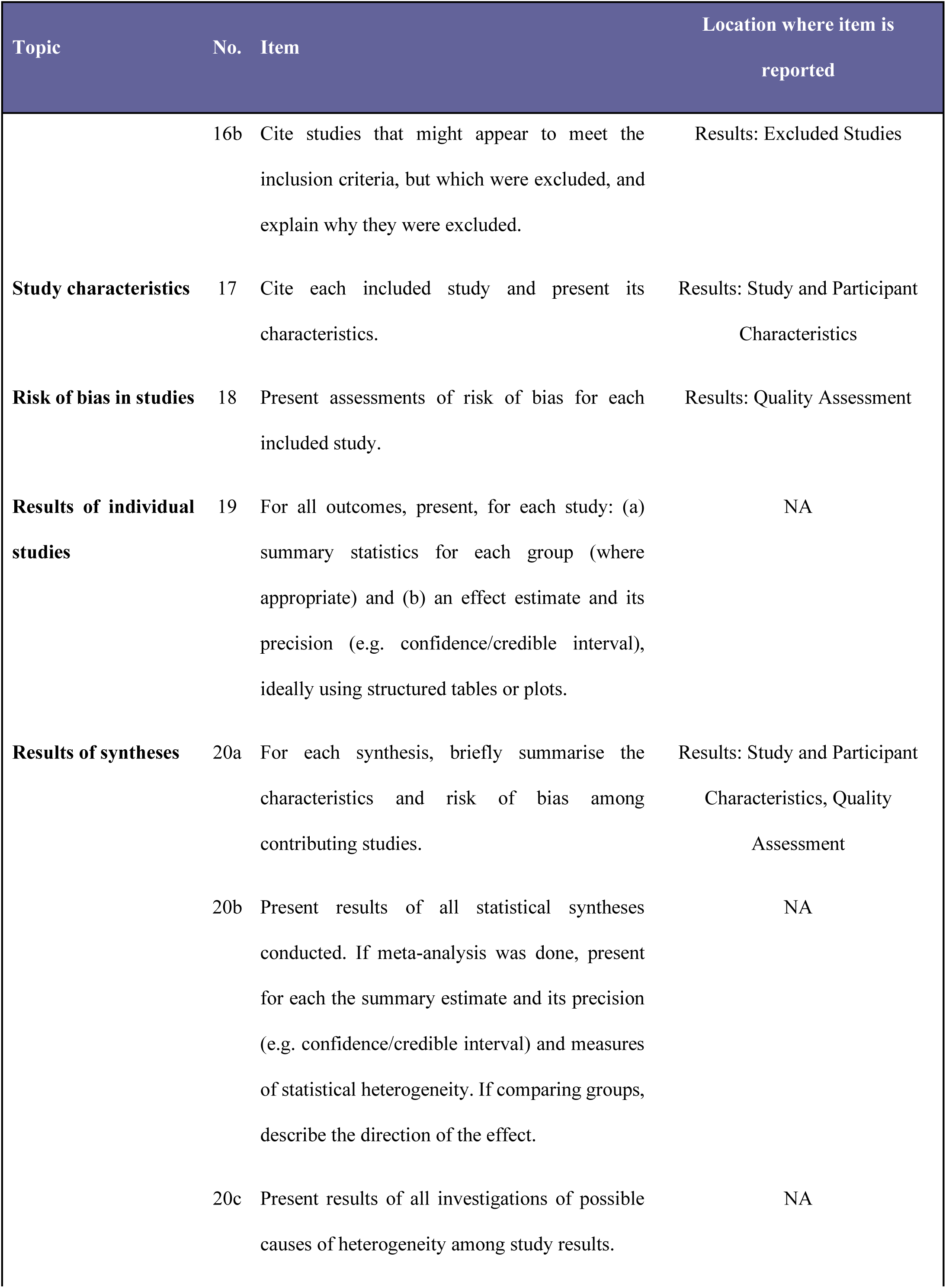
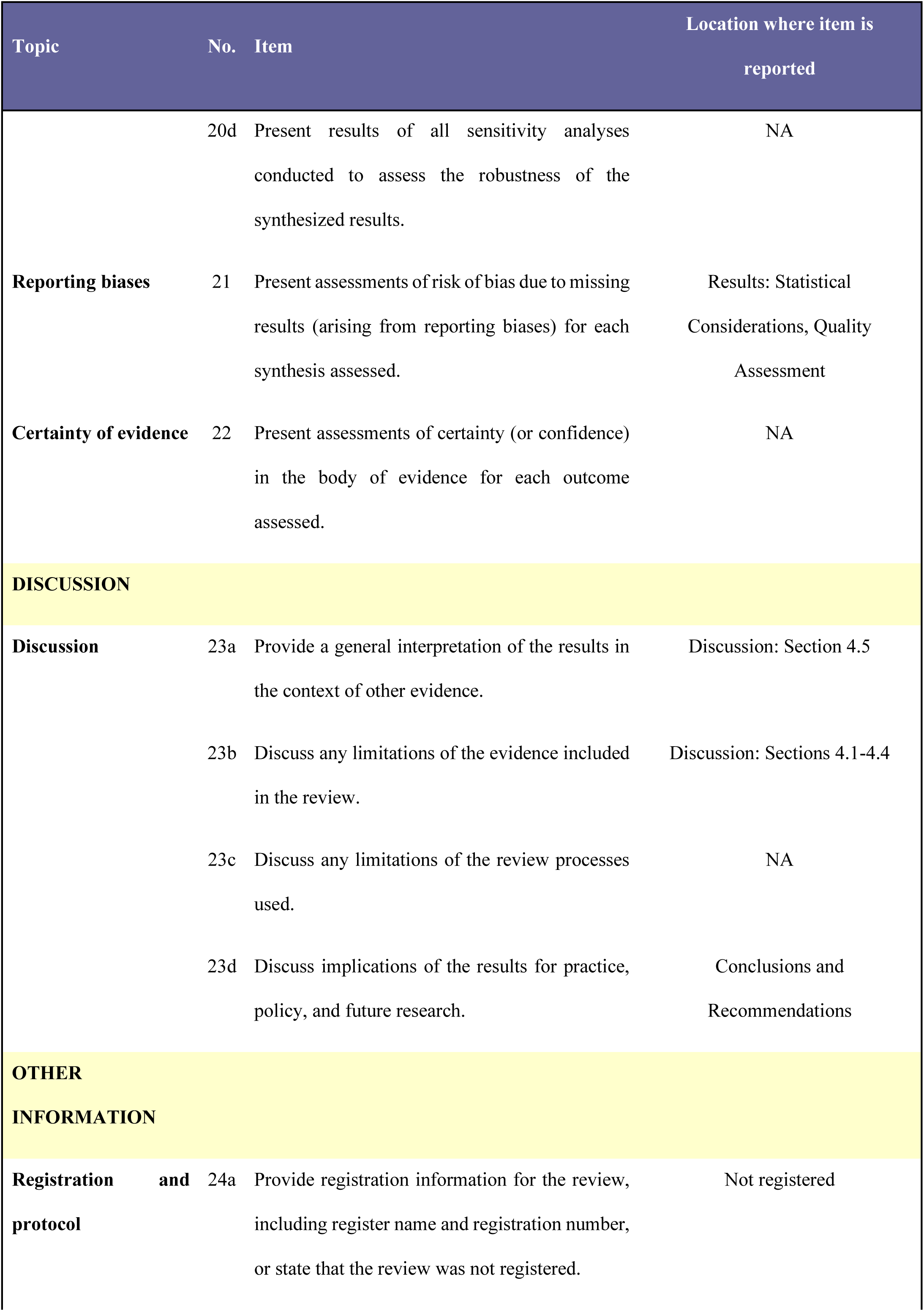
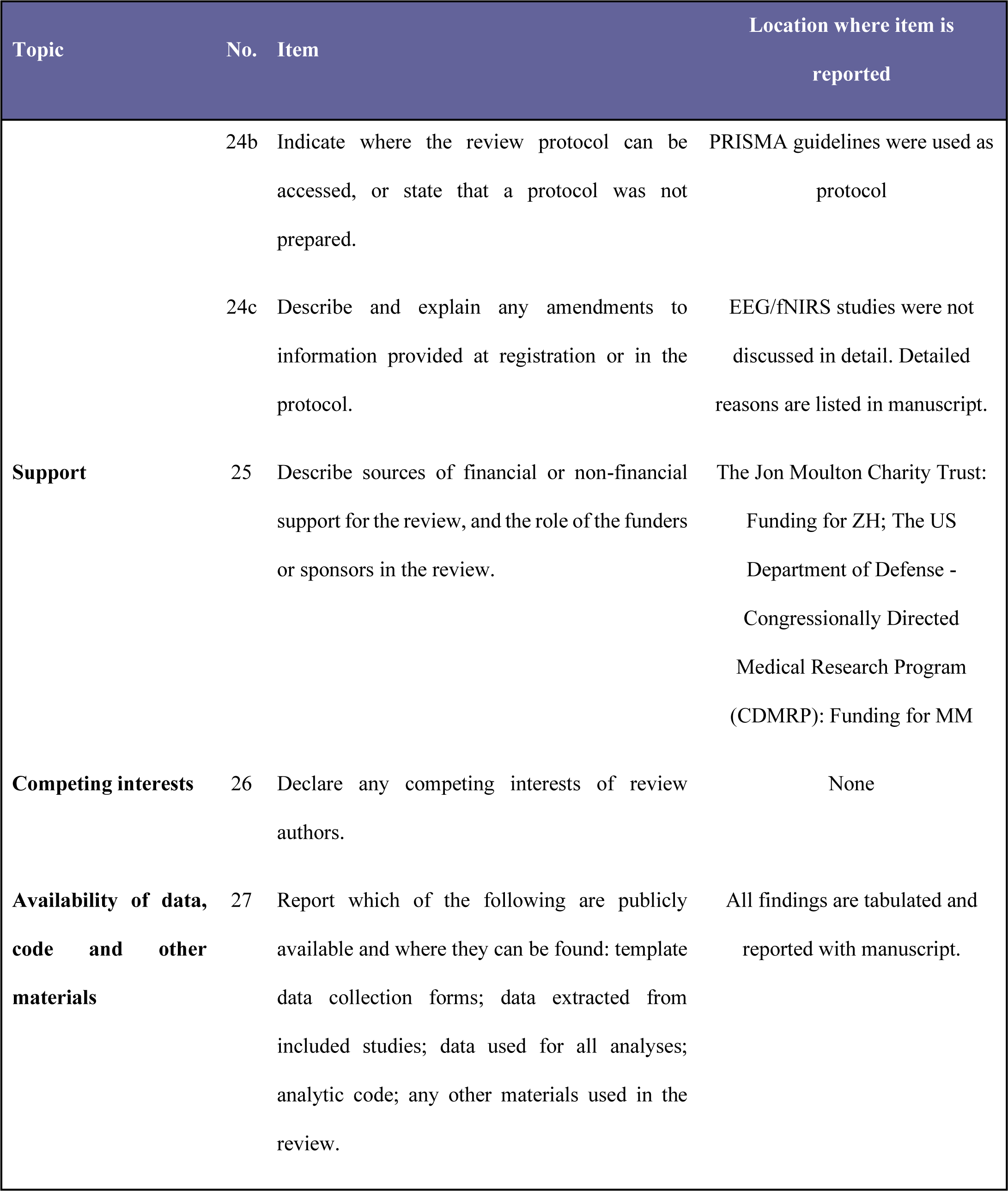
PRISMA 2020 Main Checklist.

PRIMSA Abstract Checklist.

PRISMA Abstract Checklist
| Topic | No. | Item | Reported? |
| --- | --- | --- | --- |
| **TITLE** |  |  |  |
| **Title** | 1 | Identify the report as a systematic review. | Yes |
| **BACKGROUND** |  |  |  |
| **Objectives** | 2 | Provide an explicit statement of the main objective(s) or question(s) the review addresses. | Yes |
| **METHODS** |  |  |  |
| **Eligibility criteria** | 3 | Specify the inclusion and exclusion criteria for the review. | Yes |
| **Information sources** | 4 | Specify the information sources (e.g. databases, registers) used to identify studies and the date when each was last searched. | Yes |
| **Risk of bias** | 5 | Specify the methods used to assess risk of bias in the included studies. | No |
| **Synthesis of results** | 6 | Specify the methods used to present and synthesize results. | No |
| **RESULTS** |  |  |  |
| **Included studies** | 7 | Give the total number of included studies and participants and summarise relevant characteristics of studies. | Yes |
| **Synthesis of results** | 8 | Present results for main outcomes, preferably indicating the number of included studies and participants for each. If meta-analysis was done, report the summary estimate and confidence/credible interval. If comparing groups, indicate the direction of the effect (i.e. which group is favoured). | Yes |
| **DISCUSSION** |  |  |  |
| **Limitations of evidence** | 9 | Provide a brief summary of the limitations of the evidence included in the review (e.g. study risk of bias, inconsistency and imprecision). | Yes |
| **Interpretation** | 10 | Provide a general interpretation of the results and important implications. | Yes |
| **OTHER** |  |  |  |
| **Funding** | 11 | Specify the primary source of funding for the review. | No |
| Registration | 12 | Provide the register name and registration number. | No |

## Notes

### Competing Interest Statement

The authors have declared no competing interest.

### Author Declarations

The study used previously published data as part of the systematic review.

## References

1. Adam, O., Mac Donald, C. L., Rivet, D., Ritter, J., May, T., Barefield, M., Duckworth, J., Labarge, D., Asher, D., Drinkwine, B., Woods, Y., Connor, M., & Brody, D. L. (2015). Clinical and imaging assessment of acute combat mild traumatic brain injury in Afghanistan. Neurology, 85(3), 219–227. https://doi.org/10.1212/WNL.0000000000001758

2. Aedo-Jury, F., Cottereau, B. R., Celebrini, S., & Séverac Cauquil, A. (2020). Antero-Posterior vs. Lateral Vestibular Input Processing in Human Visual Cortex. Frontiers in Integrative Neuroscience, 14, 43. https://doi.org/10.3389/FNINT.2020.00043/BIBTEX

3. Allexandre, D., Hoxha, A., Handiru, V. S., Saleh, S., Selvan, S. E., & Yue, G. H. (2019). Altered Cortical and Postural Response to Balance Perturbation in Traumatic Brain Injury - An EEG Pilot Study. Annual International Conference of the IEEE Engineering in Medicine and Biology Society. IEEE Engineering in Medicine and Biology Society. Annual International Conference, 2019, 1543–1546. https://doi.org/10.1109/EMBC.2019.8856645

4. Aravamuthan, B. R., & Angelaki, D. E. (2012). Vestibular responses in the macaque pedunculopontine nucleus and central mesencephalic reticular formation. Neuroscience, 223, 183. https://doi.org/10.1016/J.NEUROSCIENCE.2012.07.054

5. Arshad, Q., Roberts, R. E., Ahmad, H., Lobo, R., Patel, M., Ham, T., Sharp, D. J., & Seemungal, B. M. (2017). Patients with chronic dizziness following traumatic head injury typically have multiple diagnoses involving combined peripheral and central vestibular dysfunction. Clinical Neurology and Neurosurgery, 155, 17–19. https://doi.org/10.1016/J.CLINEURO.2017.01.021

6. Barmack, N. H. (2003). Central vestibular system: vestibular nuclei and posterior cerebellum. Brain Research Bulletin, 60(5–6), 511–541. https://doi.org/10.1016/S0361-9230(03)00055-8

7. Bense, S., Stephan, T., Yousry, T. A., Brandt, T., & Dieterich, M. (2001). Multisensory cortical signal increases and decreases during vestibular galvanic stimulation (fMRI). Journal of Neurophysiology, 85(2), 886–899. https://doi.org/10.1152/JN.2001.85.2.886/ASSET/IMAGES/LARGE/9K0211494006.JP EG

8. Berman, J. M., & Fredrickson, J. M. (1978). Vertigo after head injury--a five year follow-up. The Journal of Otolaryngology, 7(3), 237–245. https://europepmc.org/article/med/151151

9. Bharti, K., Suppa, A., Pietracupa, S., Upadhyay, N., Giannì, C., Leodori, G., Di Biasio, F., Modugno, N., Petsas, N., Grillea, G., Zampogna, A., Berardelli, A., & Pantano, P. (2019). Abnormal Cerebellar Connectivity Patterns in Patients with Parkinson’s Disease and Freezing of Gait. Cerebellum, 18(3), 298–308. https://doi.org/10.1007/S12311-018-0988-4/FIGURES/6

10. Bittencourt, M., van der Horn, H. J., Balart-Sánchez, S. A., Marsman, J. B. C., van der Naalt, J., & Maurits, N. M. (2022). Effects of Mild Traumatic Brain Injury on Resting State Brain Network Connectivity in Older Adults. Brain Imaging and Behavior, 16(4), 1863. https://doi.org/10.1007/S11682-022-00662-5

11. Bohnen, N. I., & Albin, R. L. (2009). Cholinergic denervation occurs early in Parkinson disease. Neurology, 73(4), 256–257. https://doi.org/10.1212/WNL.0B013E3181B0BD3D

12. Bohnen, N. I., Müller, M. L. T. M., Kotagal, V., Koeppe, R. A., Kilbourn, M. R., Gilman, S., Albin, R. L., & Frey, K. A. (2012). Heterogeneity of cholinergic denervation in Parkinson’s disease without dementia. Journal of Cerebral Blood Flow and Metabolism : Official Journal of the International Society of Cerebral Blood Flow and Metabolism, 32(8), 1609–1617. https://doi.org/10.1038/JCBFM.2012.60

13. Bohnen, N. I., Roytman, S., Griggs, A., David, S. M., Beaulieu, M. L., & Müller, M. L. T. M. (2022). Decreased vestibular efficacy contributes to abnormal balance in Parkinson’s disease. Journal of the Neurological Sciences, 440, 120357. https://doi.org/10.1016/J.JNS.2022.120357

14. Caeyenberghs, K., Leemans, A., De Decker, C., Heitger, M., Drijkoningen, D., Vander Linden, C., Sunaert, S., & Swinnen, S. P. (2012). Brain connectivity and postural control in young traumatic brain injury patients: A diffusion MRI based network analysis. NeuroImage. Clinical, 1(1), 106–115. https://doi.org/10.1016/J.NICL.2012.09.011

15. Caeyenberghs, K., Leemans, A., Geurts, M., Linden, C. Vander, Smits-Engelsman, B. C. M., Sunaert, S., & Swinnen, S. P. (2011). Correlations between white matter integrity and motor function in traumatic brain injury patients. Neurorehabilitation and Neural Repair, 25(6), 492–502. https://doi.org/10.1177/1545968310394870

16. Caeyenberghs, K., Leemans, A., Geurts, M., Taymans, T., Linden, C. Vander Smits-Engelsman, B. C. M., Sunaert, S., & Swinnen, S. P. (2010). Brain-behavior relationships in young traumatic brain injury patients: DTI metrics are highly correlated with postural control. Human Brain Mapping, 31(7), 992–1002. https://doi.org/10.1002/HBM.20911

17. Caeyenberghs, K., Siugzdaite, R., Drijkoningen, D., Marinazzo, D., & Swinnen, S. P. (2015). Functional Connectivity Density and Balance in Young Patients with Traumatic Axonal Injury. Brain Connectivity, 5(7), 423–432. https://doi.org/10.1089/BRAIN.2014.0293

18. Cai, J., Lee, S., Ba, F., Garg, S., Kim, L. J., Liu, A., Kim, D., Wang, Z. J., & McKeown, M. J. (2018). Galvanic Vestibular Stimulation (GVS) Augments Deficient Pedunculopontine Nucleus (PPN) Connectivity in Mild Parkinson’s Disease: fMRI Effects of Different Stimuli. Frontiers in Neuroscience, 12, 101. https://doi.org/10.3389/FNINS.2018.00101/BIBTEX

19. Calzolari, E., Chepisheva, M., Smith, R. M., Mahmud, M., Hellyer, P. J., Tahtis, V., Arshad, Q., Jolly, A., Wilson, M., Rust, H., Sharp, D. J., & Seemungal, B. M. (2021). Vestibular agnosia in traumatic brain injury and its link to imbalance. Brain : A Journal of Neurology, 144(1), 128–143. https://doi.org/10.1093/brain/awaa386

20. Chamelian, L., & Feinstein, A. (2004). Outcome after mild to moderate traumatic brain injury: The role of dizziness. Archives of Physical Medicine and Rehabilitation, 85(10), 1662– 1666. https://doi.org/10.1016/j.apmr.2004.02.012

21. Charney, M. F., Howell, D. R., Lanois, C., Starr, T. C., Liao, H., Coello, E., Breedlove, K. M., Meehan, W. P., Koerte, I., & Lin, A. P. (2020). Associations between Neurochemistry and Gait Performance following Concussion in Collegiate Athletes. Journal of Head Trauma Rehabilitation, 35(5), 342–353. https://doi.org/10.1097/HTR.0000000000000616

22. Churchill, N., Hutchison, M., Richards, D., Leung, G., Graham, S., & Schweizer, T. A. (2017). Brain Structure and Function Associated with a History of Sport Concussion: A Multi-Modal Magnetic Resonance Imaging Study. Journal of Neurotrauma, 34(4), 765–771. https://doi.org/10.1089/NEU.2016.4531/ASSET/IMAGES/LARGE/FIGURE2.JPEG

23. Churchill, N. W., Hutchison, M. G., Graham, S. J., & Schweizer, T. A. (2021). Acute and Chronic Effects of Multiple Concussions on Midline Brain Structures. Neurology, 97(12), e1170–e1181. https://doi.org/10.1212/WNL.0000000000012580

24. Conrad, J., Habs, M., Ruehl, R. M., Boegle, R., Ertl, M., Kirsch, V., Eren, O. E., Becker-Bense, S., Stephan, T., Wollenweber, F. A., Duering, M., Dieterich, M., & zu Eulenburg, P. (2022). Reorganization of sensory networks after subcortical vestibular infarcts: A longitudinal symptom-related voxel-based morphometry study. European Journal of Neurology, 29(5), 1514–1523. https://doi.org/10.1111/ENE.15263

25. Delano-Wood, L., Bangen, K. J., Sorg, S. F., Clark, A. L., Schiehser, D. M., Luc, N., Bondi, M. W., Werhane, M., Kim, R. T., & Bigler, E. D. (2015). Brainstem white matter integrity is related to loss of consciousness and postconcussive symptomatology in veterans with chronic mild to moderate traumatic brain injury. Brain Imaging and Behavior, 9(3), 500–512. https://doi.org/10.1007/S11682-015-9432-2

26. Della-Justina, H. M., Gamba, H. R., Lukasova, K., Nucci-da-Silva, M. P., Winkler, A. M., & Amaro, E. (2014). Interaction of brain areas of visual and vestibular simultaneous activity with fMRI. Experimental Brain Research, 233(1), 237–252. https://doi.org/10.1007/s00221-014-4107-6

27. Devilbiss, D. M., Etnoyer-Slaski, J. L., Dunn, E., Dussourd, C. R., Kothare, M. V., Martino, S. J., & Simon, A. J. (2019). Effects of Exercise on EEG Activity and Standard Tools Used to Assess Concussion. Journal of Healthcare Engineering, 2019. https://doi.org/10.1155/2019/4794637

28. Dieterich, M., Bauermann, T., Best, C., Stoeter, P., & Schlindwein, P. (2007). Evidence for cortical visual substitution of chronic bilateral vestibular failure (an fMRI study). Brain, 130(8), 2108–2116. https://doi.org/10.1093/BRAIN/AWM130

29. Dieterich, M., & Brandt, T. (2008). Functional brain imaging of peripheral and central vestibular disorders. Brain, 131(10), 2538–2552. https://doi.org/10.1093/BRAIN/AWN042

30. Diez, I., Drijkoningen, D., Stramaglia, S., Bonifazi, P., Marinazzo, D., Gooijers, J., Swinnen, S. P., & Cortes, J. M. (2017). Enhanced prefrontal functional–structural networks to support postural control deficits after traumatic brain injury in a pediatric population. Network Neuroscience, 1(2), 116. https://doi.org/10.1162/NETN_A_00007

31. Dijkstra, B. W., Bekkers, E. M. J., Gilat, M., de Rond, V., Hardwick, R. M., & Nieuwboer, A. (2020). Functional neuroimaging of human postural control: A systematic review with meta-analysis. Neuroscience & Biobehavioral Reviews, 115, 351–362. https://doi.org/10.1016/J.NEUBIOREV.2020.04.028

32. Drijkoningen, D., Caeyenberghs, K., Leunissen, I., Vander Linden, C., Sunaert, S., Duysens, J., & Swinnen, S. P. (2015). Training-induced improvements in postural control are accompanied by alterations in cerebellar white matter in brain injured patients. NeuroImage. Clinical, 7, 240–251. https://doi.org/10.1016/J.NICL.2014.12.006

33. Drijkoningen, D., Leunissen, I., Caeyenberghs, K., Hoogkamer, W., Sunaert, S., Duysens, J., & Swinnen, S. P. (2015). Regional volumes in brain stem and cerebellum are associated with postural impairments in young brain-injured patients. Human Brain Mapping, 36(12), 4897–4909. https://doi.org/10.1002/HBM.22958

34. Eickhoff, S. B., Weiss, P. H., Amunts, K., Fink, G. R., & Zilles, K. (2006). Identifying human parieto-insular vestibular cortex using fMRI and cytoarchitectonic mapping. Human Brain Mapping, 27(7), 611–621. https://doi.org/10.1002/HBM.20205

35. Elser, H., Gottesman, R. F., Walter, A. E., Coresh, J., Diaz-Arrastia, R., Mosley, T. H., & Schneider, A. L. C. (2023). Head Injury and Long-term Mortality Risk in Community-Dwelling Adults. JAMA Neurology, 80(3), 260–269. https://doi.org/10.1001/JAMANEUROL.2022.5024

36. Gera, G., Fling, B. W., & Horak, F. B. (2020). Cerebellar White Matter Damage Is Associated With Postural Sway Deficits in People With Multiple Sclerosis. Archives of Physical Medicine and Rehabilitation, 101(2), 258–264. https://doi.org/10.1016/J.APMR.2019.07.011

37. Ghajari, M., Hellyer, P. J., & Sharp, D. J. (2017). Computational modelling of traumatic brain injury predicts the location of chronic traumatic encephalopathy pathology. Brain, 140(2), 333–343. https://doi.org/10.1093/brain/aww317

38. Gordon, C. R., Levite, R., Joffe, V., & Gadoth, N. (2004). Is Posttraumatic Benign Paroxysmal Positional Vertigo Different From the Idiopathic Form? Archives of Neurology, 61(10), 1590–1593. https://doi.org/10.1001/ARCHNEUR.61.10.1590

39. Haddaway, N. R., Page, M. J., Pritchard, C. C., & McGuinness, L. A. (2022). PRISMA2020: An R package and Shiny app for producing PRISMA 2020-compliant flow diagrams, with interactivity for optimised digital transparency and Open Synthesis. Campbell Systematic Reviews, 18(2), e1230. https://doi.org/10.1002/CL2.1230

40. Hadi, Z., Mahmud, M., Pondeca, Y., Calzolari, E., Chepisheva, M., Smith, R. M., Rust, H. M., Sharp, D. J., & Seemungal, B. M. (2022). The human brain networks mediating the vestibular sensation of self-motion. Journal of the Neurological Sciences, 443. https://doi.org/10.1016/J.JNS.2022.120458

41. Hammeke, T. A., McCrea, M., Coats, S. M., Verber, M. D., Durgerian, S., Flora, K., Olsen, G. S., Leo, P. D., Gennarelli, T. A., & Rao, S. M. (2013). Acute and subacute changes in neural activation during the recovery from sport-related concussion. Journal of the International Neuropsychological Society : JINS, 19(8), 863–872. https://doi.org/10.1017/S1355617713000702

42. Handiru, V. S., Alivar, A., Hoxha, A., Saleh, S., Suviseshamuthu, E. S., Yue, G. H., & Allexandre, D. (2021). Graph-theoretical analysis of EEG functional connectivity during balance perturbation in traumatic brain injury: A pilot study. Human Brain Mapping, 42(14), 4427–4447. https://doi.org/10.1002/HBM.25554

43. Handiru, V. S., Suviseshamuthu, E. S., Saleh, S., Yue, G. H., & Allexandre, D. (2022). Identifying the Neural Correlates of Balance Deficits in Traumatic Brain Injury using the Partial Least Squares Correlation (PLSC) analysis. BioRxiv, 2022.05.15.491997. https://doi.org/10.1101/2022.05.15.491997

44. Harrell, R., Manetta, C., Guthrie, M., & Enam, N. (2023). The Prevalence of Symptom Reporting for Benign Paroxysmal Positional Vertigo in a Traumatic Brain Injury Population. Otology and Neurotology, 44(2), 172–176. https://doi.org/10.1097/MAO.0000000000003770

45. Helmchen, C., Machner, B., Rother, M., Spliethoff, P., Göttlich, M., & Sprenger, A. (2020). Effects of galvanic vestibular stimulation on resting state brain activity in patients with bilateral vestibulopathy. Human Brain Mapping, 41(9), 2527–2547. https://doi.org/10.1002/HBM.24963

46. Helmchen, C., Ye, Z., Sprenger, A., & Münte, T. F. (2014). Changes in resting-state fMRI in vestibular neuritis. Brain Structure and Function, 219(6), 1889–1900. https://doi.org/10.1007/s00429-013-0608-5

47. Helmich, I., Berger, A., & Lausberg, H. (2016). Neural Control of Posture in Individuals with Persisting Postconcussion Symptoms. Medicine and Science in Sports and Exercise, 48(12), 2362–2369. https://doi.org/10.1249/MSS.0000000000001028

48. Helmich, I., Coenen, J., Henckert, S., Pardalis, E., Schupp, S., & Lausberg, H. (2020). Reduced frontopolar brain activation characterizes concussed athletes with balance deficits. NeuroImage: Clinical, 25, 102164. https://doi.org/10.1016/J.NICL.2020.102164

49. Hou, J., Mohanty, R., Chu, D., Nair, V. A., Danilov, Y., Kaczmarek, K. A., Meyerand, B., Tyler, M., & Prabhakaran, V. (2022). Translingual neural stimulation affects resting-state functional connectivity in mild-moderate traumatic brain injury. Journal of Neuroimaging, 32(6), 1193. https://doi.org/10.1111/JON.13029

50. Howell, D. R., Meehan, W. P., Barber Foss, K. D., Reches, A., Weiss, M., & Myer, G. D. (2018). Reduced dual-task gait speed is associated with visual Go/No-Go brain network activation in children and adolescents with concussion. https://doi.org/10.1080/02699052.2018.1482424, 32(9), 1129–1134. https://doi.org/10.1080/02699052.2018.1482424

51. Jacob, D., Unnsteinsdóttir Kristensen, I. S., Aubonnet, R., Recenti, M., Donisi, L., Ricciardi, C., Svansson, H. Á. R., Agnarsdóttir, S., Colacino, A., Jónsdóttir, M. K., Kristjánsdóttir, H., Sigurjónsdóttir, H., Cesarelli, M., Eggertsdóttir Claessen, L. Ó., Hassan, M., Petersen, H., & Gargiulo, P. (2022). Towards defining biomarkers to evaluate concussions using virtual reality and a moving platform (BioVRSea). Scientific Reports, 12(1). https://doi.org/10.1038/S41598-022-12822-0

52. Jahn, K., Deutschländer, A., Stephan, T., Strupp, M., Wiesmann, M., & Brandt, T. (2004). Brain activation patterns during imagined stance and locomotion in functional magnetic resonance imaging. NeuroImage, 22(4), 1722–1731. https://doi.org/10.1016/J.NEUROIMAGE.2004.05.017

53. Jang, S. H., Kim, T. H., Kwon, Y. H., Lee, M. Y., & Lee, H. Do. (2016). Postural Instability in Patients With Injury of Corticoreticular Pathway Following Mild Traumatic Brain Injury. American Journal of Physical Medicine & Rehabilitation, 95(8), 580–587. https://doi.org/10.1097/PHM.0000000000000446

54. Jang, S. H., Yi, J. H., & Kwon, H. G. (2016). Injury of the inferior cerebellar peduncle in patients with mild traumatic brain injury: A diffusion tensor tractography study. Brain Injury, 30(10), 1271–1275. https://doi.org/10.1080/02699052.2016.1178805

55. Jolly, A. E., Bălăeţ, M., Azor, A., Friedland, D., Sandrone, S., Graham, N. S. N., Zimmerman, K., & Sharp, D. J. (2020). Detecting axonal injury in individual patients after traumatic brain injury. Brain. https://doi.org/10.1093/brain/awaa372

56. Joubran, K., Bar-Haim, S., & Shmuelof, L. (2022). The functional and structural neural correlates of dynamic balance impairment and recovery in persons with acquired brain injury. Scientific Reports 2022 12:1, 12(1), 1–11. https://doi.org/10.1038/s41598-022-12123-6

57. Kahane, P., Hoffmann, D., Minotti, L., & Berthoz, A. (2003). Reappraisal of the Human Vestibular Cortex by Cortical Electrical Stimulation Study. Annals of Neurology, 54(5), 615–624. https://doi.org/10.1002/ana.10726

58. Karim, H. T., Sparto, P. J., Aizenstein, H. J., Furman, J. M., Huppert, T. J., Erickson, K. I., & Loughlin, P. J. (2014). Functional MR imaging of a simulated balance task. Brain Research, 1555, 20–27. https://doi.org/10.1016/J.BRAINRES.2014.01.033

59. Kaushal, M., España, L. Y., Nencka, A. S., Wang, Y., Nelson, L. D., McCrea, M. A., & Meier, T. B. (2019). Resting-state functional connectivity after concussion is associated with clinical recovery. Human Brain Mapping, 40(4), 1211–1220. https://doi.org/10.1002/HBM.24440

60. Kheradmand, A., & Zee, D. S. (2011). Cerebellum and Ocular Motor Control. Frontiers in Neurology, 2. https://doi.org/10.3389/FNEUR.2011.00053

61. Kim, E., Seo, H. G., Lee, H. H., Lee, S. H., Choi, S. H., Yoo, R. E., Cho, W. S., Wagner, A. K., & Oh, B. M. (2019). Altered white matter integrity after mild to moderate traumatic brain injury. Journal of Clinical Medicine, 8(9). https://doi.org/10.3390/jcm8091318

62. Kim, S. Y., Park, J. E., Lee, Y. J., Seo, H. J., Sheen, S. S., Hahn, S., Jang, B. H., & Son, H. J. (2013). Testing a tool for assessing the risk of bias for nonrandomized studies showed moderate reliability and promising validity. Journal of Clinical Epidemiology, 66(4), 408–414. https://doi.org/10.1016/J.JCLINEPI.2012.09.016

63. Kirsch, V., Keeser, D., Hergenroeder, T., Erat, O., Ertl-Wagner, B., Brandt, T., & Dieterich, M. (2016). Structural and functional connectivity mapping of the vestibular circuitry from human brainstem to cortex. Brain Structure and Function, 221(3), 1291–1308. https://doi.org/10.1007/s00429-014-0971-x

64. Lancaster, M. A., Meier, T. B., Olson, D. V., McCrea, M. A., Nelson, L. D., & Muftuler, L. T. (2018). Chronic differences in white matter integrity following sport-related concussion as measured by diffusion MRI: 6-Month follow-up. Human Brain Mapping, 39(11), 4276–4289. https://doi.org/10.1002/HBM.24245

65. Langlois, J. A., Rutland-Brown, W., & Wald, M. M. (2006). The epidemiology and impact of traumatic brain injury: A brief overview. Journal of Head Trauma Rehabilitation, 21(5), 375–378. https://doi.org/10.1097/00001199-200609000-00001

66. Lempke, L. B., Hoch, M. C., Call, J. A., Schmidt, J. D., & Lynall, R. C. (2023). Lower Extremity Somatosensory Function Throughout Concussion Recovery: A Prospective Cohort Study. Journal of Head Trauma Rehabilitation, 38(2), E156–E166. https://doi.org/10.1097/HTR.0000000000000805

67. Liang, X., Yeh, C. H., Domínguez D., J. F., Poudel, G., Swinnen, S. P., & Caeyenberghs, K. (2021). Longitudinal fixel-based analysis reveals restoration of white matter alterations following balance training in young brain-injured patients. NeuroImage: Clinical, 30, 102621. https://doi.org/10.1016/j.nicl.2021.102621

68. Lobel, E., Kleine, J. F., Le Bihan, D., Leroy-Willig, A., & Berthoz, A. (1998). Functional MRI of galvanic vestibular stimulation. Journal of Neurophysiology, 80(5), 2699–2709. https://doi.org/10.1152/JN.1998.80.5.2699/ASSET/IMAGES/LARGE/JNP.NO01F4.JPEG

69. Maas, A. I. R., Menon, D. K., David Adelson, P. D., Andelic, N., Bell, M. J., Belli, A., Bragge, P., Brazinova, A., Büki, A., Chesnut, R. M., Citerio, G., Coburn, M., Jamie Cooper, D., Tamara Crowder, A., Czeiter, E., Czosnyka, M., Diaz-Arrastia, R., Dreier, J. P., Duhaime, A. C., … Zemek, R. (2017). Traumatic brain injury: integrated approaches to improve prevention, clinical care, and research. The Lancet Neurology, 16(12), 987–1048. https://doi.org/10.1016/S1474-4422(17)30371-X

70. Maas, A. I. R., Menon, D. K., Manley, G. T., Abrams, M., Åkerlund, C., Andelic, N., Aries, M., Bashford, T., Bell, M. J., Bodien, Y. G., Brett, B. L., Büki, A., Chesnut, R. M., Citerio, G., Clark, D., Clasby, B., Cooper, D. J., Czeiter, E., Czosnyka, M., … Zemek, R. (2022). Traumatic brain injury: progress and challenges in prevention, clinical care, and research. The Lancet Neurology, 21(11), 1004–1060. https://doi.org/10.1016/S1474-4422(22)00309-X/ATTACHMENT/21111770-6322-463D-BB3A-652E385F959D/MMC1.PDF

71. Madaan, P., Gupta, D., Agrawal, D., Kumar, A., Jauhari, P., Chakrabarty, B., Sharma, S., Pandey, R. M., Paul, V. K., Misra, M. C., & Gulati, S. (2021). Neurocognitive Outcomes and Their Diffusion Tensor Imaging Correlates in Children With Mild Traumatic Brain Injury. Journal of Child Neurology, 36(8), 664–672. https://doi.org/10.1177/0883073821996095

72. Madhavan, R., Joel, S. E., Mullick, R., Cogsil, T., Niogi, S. N., Tsiouris, A. J., Mukherjee, P., Masdeu, J. C., Marinelli, L., & Shetty, T. (2019). Longitudinal Resting State Functional Connectivity Predicts Clinical Outcome in Mild Traumatic Brain Injury. Journal of Neurotrauma, 36(5), 650–660. https://doi.org/10.1089/NEU.2018.5739

73. Marcus, H. J., Paine, H., Sargeant, M., Wolstenholme, S., Collins, K., Marroney, N., Arshad, Q., Tsang, K., Jones, B., Smith, R., Wilson, M. H., Rust, H. M., & Seemungal, B. M. (2019). Vestibular dysfunction in acute traumatic brain injury. Journal of Neurology. https://doi.org/10.1007/s00415-019-09403-z

74. Marek, S., Tervo-Clemmens, B., Calabro, F. J., Montez, D. F., Kay, B. P., Hatoum, A. S., Donohue, M. R., Foran, W., Miller, R. L., Hendrickson, T. J., Malone, S. M., Kandala, S., Feczko, E., Miranda-Dominguez, O., Graham, A. M., Earl, E. A., Perrone, A. J., Cordova, M., Doyle, O., … Dosenbach, N. U. F. (2022). Reproducible brain-wide association studies require thousands of individuals. Nature 2022 603:7902, 603(7902), 654–660. https://doi.org/10.1038/s41586-022-04492-9

75. Maskell, F., Chiarelli, P., & Isles, R. (2006). Dizziness after traumatic brain injury: Overview and measurement in the clinical setting. Brain Injury, 20(3), 293–305. https://doi.org/10.1080/02699050500488041

76. McGuinness, L. A., & Higgins, J. P. T. (2021). Risk-of-bias VISualization (robvis): An R package and Shiny web app for visualizing risk-of-bias assessments. Research Synthesis Methods, 12(1), 55–61. https://doi.org/10.1002/JRSM.1411

77. Meier, T. B., Giraldo-Chica, M., España, L. Y., Mayer, A. R., Harezlak, J., Nencka, A. S., Wang, Y., Koch, K. M., Wu, Y. C., Saykin, A. J., Giza, C. C., Goldman, J., Difiori, J. P., Guskiewicz, K. M., Mihalik, J. P., Brooks, A., Broglio, S. P., McAllister, T., & McCrea, M. A. (2020). Resting-State fMRI Metrics in Acute Sport-Related Concussion and Their Association with Clinical Recovery: A Study from the NCAA-DOD CARE Consortium. Journal of Neurotrauma, 37(1), 152–162. https://doi.org/10.1089/NEU.2019.6471

78. Mitsutake, T., Sakamoto, M., Kawaguchi, A., Tamari, M., & Horikawa, E. (2020). Greater functional activation during galvanic vestibular stimulation is associated with improved postural stability: a GVS-fMRI study. https://doi.org/10.1080/08990220.2020.1803256, 37(4), 257–261. https://doi.org/10.1080/08990220.2020.1803256

79. Muftuler, L. T., Meier, T. B., Keith, M., Budde, M. D., Huber, D. L., & McCrea, M. A. (2020). Serial Diffusion Kurtosis Magnetic Resonance Imaging Study during Acute, Subacute, and Recovery Periods after Sport-Related Concussion. Journal of Neurotrauma, 37(19), 2081–2092. https://doi.org/10.1089/NEU.2020.6993

80. Müller, M. L. T. M., Albin, R. L., Kotagal, V., Koeppe, R. A., Scott, P. J. H., Frey, K. A., & Bohnen, N. I. (2013). Thalamic cholinergic innervation and postural sensory integration function in Parkinson’s disease. Brain, 136(11), 3282–3289. https://doi.org/10.1093/BRAIN/AWT247

81. Nigmatullina, Y., Hellyer, P. J., Nachev, P., Sharp, D. J., & Seemungal, B. M. (2015). The neuroanatomical correlates of training-related perceptuo-reflex uncoupling in dancers. Cerebral Cortex, 25(2), 554–562. https://doi.org/10.1093/cercor/bht266

82. Odom, A. D., Richmond, S. B., & Fling, B. W. (2021). White Matter Microstructure of the Cerebellar Peduncles Is Associated with Balance Performance during Sensory Re-Weighting in People with Multiple Sclerosis. Cerebellum, 20(1), 92–100. https://doi.org/10.1007/S12311-020-01190-Y/FIGURES/3

83. Oka, S., Ikeda, T., Mitsutake, T., Ogata, K., & Goto, Y. (2022). Unilateral cathodal transcranial direct current stimulation over the parietal area modulates on postural control depending with eyes open and closed. MedRxiv, 2022.05.16.22275178. https://doi.org/10.1101/2022.05.16.22275178

84. Ouchi, Y., Okada, H., Yoshikawa, E., Nobezawa, S., & Futatsubashi, M. (1999). Brain activation during maintenance of standing postures in humans. Brain, 122(2), 329–338. https://doi.org/10.1093/BRAIN/122.2.329

85. Page, M. J., McKenzie, J. E., Bossuyt, P. M., Boutron, I., Hoffmann, T. C., Mulrow, C. D., Shamseer, L., Tetzlaff, J. M., Akl, E. A., Brennan, S. E., Chou, R., Glanville, J., Grimshaw, J. M., Hróbjartsson, A., Lalu, M. M., Li, T., Loder, E. W., Mayo-Wilson, E., McDonald, S., … Moher, D. (2021). The PRISMA 2020 statement: an updated guideline for reporting systematic reviews. BMJ, 372. https://doi.org/10.1136/BMJ.N71

86. Piantino, J., Schwartz, D. L., Luther, M., Newgard, C., Silbert, L., Raskind, M., Pagulayan, K., Kleinhans, N., Iliff, J., & Peskind, E. (2021). Link between Mild Traumatic Brain Injury, Poor Sleep, and Magnetic Resonance Imaging: Visible Perivascular Spaces in Veterans. Journal of Neurotrauma, 38(17), 2391. https://doi.org/10.1089/NEU.2020.7447

87. Prosperini, L., Fanelli, F., Petsas, N., Sbardella, E., Tona, F., Raz, E., Fortuna, D., De Angelis, F., Pozzilli, C., & Pantano, P. (2014). Multiple Sclerosis: Changes in Microarchitecture of White Matter Tracts after Training with a Video Game Balance Board. https://doi.org/10.1148/Radiol.14140168, 273(2), 529–538. https://doi.org/10.1148/RADIOL.14140168

88. Prosperini, L., Sbardella, E., Raz, E., Cercignani, M., Tona, F., Bozzali, M., Petsas, N., Pozzilli, C., & Pantano, P. (2013). Multiple Sclerosis: White and Gray Matter Damage Associated with Balance Deficit Detected at Static Posturography. https://doi.org/10.1148/Radiol.13121695, 268(1), 181–189. https://doi.org/10.1148/RADIOL.13121695

89. Rosario, B. L., Rosso, A. L., Aizenstein, H. J., Harris, T., Newman, A. B., Satterfield, S., Studenski, S. A., Yaffe, K., & Rosano, C. (2016). Cerebral white matter and slow gait: Contribution of hyperintensities and normal-appearing parenchyma. Journals of Gerontology - Series A Biological Sciences and Medical Sciences, 71(7), 968–973. https://doi.org/10.1093/gerona/glv224

90. Rust, H. M., Smith, R. M., Mahmud, M., Golding, J. F., & Seemungal, B. M. (2022). Force dependency of benign paroxysmal positional vertigo in acute traumatic brain injury: a prospective study. Journal of Neurology, Neurosurgery & Psychiatry, jnnp-2022–328997. https://doi.org/10.1136/JNNP-2022-328997

91. Sargeant, M., Sykes, E., Saviour, M., Sawhney, A., Calzolari, E., Arthur, J., McGoldrick, A., & Seemungal, B. (2018). The utility of the Sports Concussion Assessment Tool in hospitalized traumatic brain injury patients. Journal of Concussion, 2, 205970021880812. https://doi.org/10.1177/2059700218808121

92. Schlosser, H. G., Lindemann, J. N., Vajkoczy, P., & Clarke, A. H. (2009). Vestibulo-ocular monitoring as a predictor of outcome after severe traumatic brain injury. *Critical Care (London*, England), 13(6), 1–10. https://doi.org/10.1186/CC8187/TABLES/4

93. Seemungal, B. M., & Passamonti, L. (2018). Persistent postural-perceptual dizziness: A useful new syndrome. In Practical Neurology (Vol. 18, Issue 1, pp. 3–4). BMJ Publishing Group. https://doi.org/10.1136/practneurol-2017-001817

94. Sehm, B., Taubert, M., Conde, V., Weise, D., Classen, J., Dukart, J., Draganski, B., Villringer, A., & Ragert, P. (2014). Structural brain plasticity in Parkinson’s disease induced by balance training. Neurobiology of Aging, 35(1), 232–239. https://doi.org/10.1016/J.NEUROBIOLAGING.2013.06.021

95. Seo, M.-H., Park, S.-H., Ko, M.-H., & Seo, J.-H. (2011). Motor Evoked Potentials of Trunk Muscles in Traumatic Brain Injury Patients. Annals of Rehabilitation Medicine, 35(4), 557. https://doi.org/10.5535/ARM.2011.35.4.557

96. Shetty, T., Nguyen, J. T., Cogsil, T., Tsiouris, A. J., Niogi, S. N., Kim, E. U., Dalal, A., Halvorsen, K., Cummings, K., Zhang, T., Masdeu, J. C., Mukherjee, P., & Marinelli, L. (2018). Clinical Findings in a Multicenter MRI Study of Mild TBI. Frontiers in Neurology, 9(OCT). https://doi.org/10.3389/FNEUR.2018.00836

97. Slobounov, S., Sebastianelli, W., & Hallett, M. (2012). Residual brain dysfunction observed one year post-mild traumatic brain injury: Combined EEG and balance study. Clinical Neurophysiology : Official Journal of the International Federation of Clinical Neurophysiology, 123(9), 1755. https://doi.org/10.1016/J.CLINPH.2011.12.022

98. Slobounov, S., Sebastianelli, W., & Moss, R. (2005). Alteration of posture-related cortical potentials in mild traumatic brain injury. Neuroscience Letters, 383(3), 251–255. https://doi.org/10.1016/J.NEULET.2005.04.039

99. Smith, R. M., Marroney, N., Beattie, J., Newdick, A., Tahtis, V., Burgess, C., Marsden, J., & Seemungal, B. M. (2020). A mixed methods randomised feasibility trial investigating the management of benign paroxysmal positional vertigo in acute traumatic brain injury. Pilot and Feasibility Studies, 6(1), 1–10. https://doi.org/10.1186/S40814-020-00669-Z/FIGURES/4

100. Sours, C., Zhuo, J., Roys, S., Shanmuganathan, K., & Gullapalli, R. P. (2015). Disruptions in Resting State Functional Connectivity and Cerebral Blood Flow in Mild Traumatic Brain Injury Patients. PloS One, 10(8). https://doi.org/10.1371/JOURNAL.PONE.0134019

101. Spisak, T., Bingel, U., & Wager, T. D. (2023). Multivariate BWAS can be replicable with moderate sample sizes. Nature 2023 615:7951, 615(7951), E4–E7. https://doi.org/10.1038/s41586-023-05745-x

102. Stiles, L., & Smith, P. F. (2015). The vestibular–basal ganglia connection: Balancing motor control. Brain Research, 1597, 180–188. https://doi.org/10.1016/J.BRAINRES.2014.11.063

103. Surgent, O. J., Dadalko, O. I., Pickett, K. A., & Travers, B. G. (2019). Balance and the Brain: A Review of Structural Brain Correlates of Postural Balance and Balance Training in Humans. Gait & Posture, 71, 245. https://doi.org/10.1016/J.GAITPOST.2019.05.011

104. Taubert, M., Draganski, B., Anwander, A., Müller, K., Horstmann, A., Villringer, A., & Ragert, P. (2010). Dynamic Properties of Human Brain Structure: Learning-Related Changes in Cortical Areas and Associated Fiber Connections. The Journal of Neuroscience, 30(35), 11670. https://doi.org/10.1523/JNEUROSCI.2567-10.2010

105. Teel, E. F., Ray, W. J., Geronimo, A. M., & Slobounov, S. M. (2014). Residual alterations of brain electrical activity in clinically asymptomatic concussed individuals: an EEG study. Clinical Neurophysiology : Official Journal of the International Federation of Clinical Neurophysiology, 125(4), 703–707. https://doi.org/10.1016/J.CLINPH.2013.08.027

106. Thompson, J., Sebastianelli, W., & Slobounov, S. (2005). EEG and postural correlates of mild traumatic brain injury in athletes. Neuroscience Letters, 377(3), 158–163. https://doi.org/10.1016/J.NEULET.2004.11.090

107. Toth, L., Czigler, A., Horvath, P., Kornyei, B., Szarka, N., Schwarcz, A., Ungvari, Z., Buki, A., & Toth, P. (2021). Traumatic brain injury-induced cerebral microbleeds in the elderly. GeroScience, 43(1), 125–136. https://doi.org/10.1007/S11357-020-00280-3

108. Trinidade, A., Cabreira, V., Goebel, J. A., Staab, J. P., Kaski, D., & Stone, J. (2023). Predictors of persistent postural-perceptual dizziness (PPPD) and similar forms of chronic dizziness precipitated by peripheral vestibular disorders: a systematic review. *Journal of Neurology*, Neurosurgery & Psychiatry, 0, 1–12. https://doi.org/10.1136/JNNP-2022-330196

109. Urban, K., Schudlo, L., Keightley, M., Alain, S., Reed, N., & Chau, T. (2020). Altered Brain Activation in Youth following Concussion: Using a Dual-task Paradigm. https://doi.org/10.1080/17518423.2020.1825539, 24(3), 187–198. https://doi.org/10.1080/17518423.2020.1825539

110. van der Veen, S. M., Perera, R. A., Manning-Franke, L., Agyemang, A. A., Skop, K., Sponheim, S. R., Wilde, E. A., Stamenkovic, A., Thomas, J. S., & Walker, W. C. (2023). Executive function and relation to static balance metrics in chronic mild TBI: A LIMBIC-CENC secondary analysis. Frontiers in Neurology, 13, 2651. https://doi.org/10.3389/FNEUR.2022.906661/BIBTEX

111. Vartanian, O., Coady, L., Blackler, K., Fraser, B., & Cheung, B. (2021). Neuropsychological, Neurocognitive, Vestibular, and Neuroimaging Correlates of Exposure to Repetitive Low-Level Blast Waves: Evidence From Four Nonoverlapping Samples of Canadian Breachers. Military Medicine, 186(3–4), e393–e400. https://doi.org/10.1093/MILMED/USAA332

112. Villiger, M., Grabher, P., Hepp-Reymond, M. C., Kiper, D., Curt, A., Bolliger, M., Hotz-Boendermaker, S., Kollias, S., Eng, K., & Freund, P. (2015). Relationship between structural brainstem and brain plasticity and lower-limb training in spinal cord injury: a longitudinal pilot study. Frontiers in Human Neuroscience, 9(MAY). https://doi.org/10.3389/FNHUM.2015.00254

113. Von Brevern, M., Zeise, D., Neuhauser, H., Clarke, A. H., & Lempert, T. (2005). Acute migrainous vertigo: clinical and oculographic findings. Brain : A Journal of Neurology, 128(Pt 2), 365–374. https://doi.org/10.1093/BRAIN/AWH351

114. Walter, A., Finelli, K., Bai, X., Arnett, P., Bream, T., Seidenberg, P., Lynch, S., Johnson, B., & Slobounov, S. (2017). Effect of Enzogenol® Supplementation on Cognitive, Executive, and Vestibular/Balance Functioning in Chronic Phase of Concussion. Developmental Neuropsychology, 42(2), 93–103. https://doi.org/10.1080/87565641.2016.1256404

115. Wang, Y., Nencka, A. S., Meier, T. B., Guskiewicz, K., Mihalik, J. P., Alison Brooks, M., Saykin, A. J., Koch, K. M., Wu, Y. C., Nelson, L. D., McAllister, T. W., Broglio, S. P., & McCrea, M. A. (2019). Cerebral blood flow in acute concussion: preliminary ASL findings from the NCAA-DoD CARE consortium. Brain Imaging and Behavior, 13(5), 1375–1385. https://doi.org/10.1007/S11682-018-9946-5

116. Weng, L., Xie, Q., Zhao, L., Zhang, R., Ma, Q., Wang, J., Jiang, W., He, Y., Chen, Y., Li, C., Ni, X., Xu, Q., Yu, R., & Huang, R. (2017). Abnormal structural connectivity between the basal ganglia, thalamus, and frontal cortex in patients with disorders of consciousness. Cortex; a Journal Devoted to the Study of the Nervous System and Behavior, 90, 71–87. https://doi.org/10.1016/J.CORTEX.2017.02.011

117. Wood, N. I., Hentig, J., Hager, M., Hill-Pearson, C., Hershaw, J. N., Souvignier, A. R., & Bobula, S. A. (2022). The Non-Concordance of Self-Reported and Performance-Based Measures of Vestibular Dysfunction in Military and Civilian Populations Following TBI. Journal of Clinical Medicine, 11(11), 2959. https://doi.org/10.3390/JCM11112959

118. Woytowicz, E. J., Sours, C., Gullapalli, R. P., Rosenberg, J., & Westlake, K. P. (2018). Modulation of working memory load distinguishes individuals with and without balance impairments following mild traumatic brain injury. Brain Injury, 32(2), 191–199. https://doi.org/10.1080/02699052.2017.1403045

119. Yousif, N., Bhatt, H., Bain, P. G., Nandi, D., & Seemungal, B. M. (2016). The effect of pedunculopontine nucleus deep brain stimulation on postural sway and vestibular perception. European Journal of Neurology, 23(3), 668–670. https://doi.org/10.1111/ene.12947

120. Zwergal, A., Linn, J., Xiong, G., Brandt, T., Strupp, M., & Jahn, K. (2012). Aging of human supraspinal locomotor and postural control in fMRI. Neurobiology of Aging, 33(6), 1073– 1084. https://doi.org/10.1016/J.NEUROBIOLAGING.2010.09.022

